# A Randomised, Triple-Blind, Dose-Finding Study of the Impact of Psilocybin on Motor Function in Healthy Participants

**DOI:** 10.64898/2025.12.22.25342874

**Authors:** Chiranth Bhagavan, Olivia Carter, Glenn Nielsen, David Berlowitz, Sara Issak, Sabine Braat, Sophie Zaloumis, Zachary Attard, Gina Oliver, Deanne Mayne, James Rucker, Matthew Butler, Orwa Dandash, Alexander Bryson, Richard A. Kanaan

**Affiliations:** Department of Psychiatry, University of Melbourne, Austin Health, Heidelberg, VIC, Australia; Melbourne School of Psychological Sciences, University of Melbourne, Parkville, VIC, Australia; Neurosciences, City St George’s, University of London, London, United Kingdom; Institute for Breathing and Sleep, Austin Health, Heidelberg, VIC, Australia; Department of Physiotherapy, Melbourne School of Health Sciences, The University of Melbourne, Carlton, VIC, Australia; Department of Physiotherapy, Epworth Healthcare, Camberwell, VIC, Australia; Centre for Epidemiology and Biostatistics, Melbourne School of Population and Global Health, The University of Melbourne, Melbourne, VIC, Australia; MISCH (Methods and Implementation Support for Clinical Health) Research Hub, Faculty of Medicine, Dentistry and Health Sciences, University of Melbourne, Melbourne, VIC, Australia; FND Hope International, Salmon, Idaho, United States; Department of Psychological Medicine, Institute of Psychiatry, Psychology & Neuroscience, King’s College London, London, United Kingdom; South London & Maudsley NHS Foundation Trust, Bethlem Royal Hospital, Beckenham, United Kingdom; Florey Institute of Neuroscience & Mental Health, Parkville, VIC, Australia; Department of Neurology, Melbourne Health, Parkville, VIC, Australia; Department of Neurology, Eastern Health, Box Hill, VIC, Australia

**Author notes:** Contributed equally to the paper (Alexander Bryson, Richard A. Kanaan). Corresponding author: Dr Chiranth Bhagavan Department of Psychiatry, University of Melbourne, Austin Health, 145 Studley Rd, Heidelberg VIC 3084, Australia.

**Keywords:** Psilocybin, psychedelics, motor function, physiotherapy, rehabilitation

## Abstract

**Background:** Psychedelics exert widespread effects on brain activity, but their impact on motor function is unclear. This is clinically relevant given the emerging interest in psychedelic-assisted physical therapy for disorders of motor function. This study’s primary objectives examined the feasibility and safety of administering movement tasks following low-to-moderate doses of psilocybin in healthy volunteers.

**Methods:** Healthy participants were randomly assigned three psilocybin doses consisting of either (1) 5mg, 10mg, and 15mg, or (2) 10mg, 15mg, and 20mg, with at least one week between doses. Movement tasks were administered during the acute drug effects. Participants, physiotherapists, and statisticians were blinded to the dosing order. Feasibility was assessed by evaluating completion of the de Morton Mobility Index and Functional Movement Exploration (measures of gross motor function). Safety outcomes included vital signs and adverse events. Additional exploratory motor outcomes included the Action Research Arm Test (assessing dexterity), Box and Block Test (Original and Modified versions) (combining dexterity with motor speed), Digit Symbol Substitution Test (combining motor speed with intellectual functions), and Reaction Time Ruler Drop Test (assessing reaction time). The 5-Dimensional Altered States of Consciousness and Ego-Dissolution Inventory assessed changes in states of consciousness. Blinding efficacy was assessed by asking participants and physiotherapists to guess the doses administered.

**Results:** Thirteen participants were randomised: seven to 5mg, 10mg, and 15mg; six to 10mg, 15mg, and 20mg. One participant was unable to complete several movement tasks at 20mg. Nausea (n=8, 62%) and headache (n=7, 54%) were the most common adverse events. No serious adverse events or adverse events related to movement task administration occurred. Median values [interquartile ranges] remained near-perfect across doses for the de Morton Mobility Index (92.5-100.0 [85.0-100.0]), Functional Movement Exploration (100.0 [96.0-100.0]), and Action Research Arm Test (56.0-57.0 [52.0-57.0]). Baseline Box and Block Test (Original) median scores (65.0 [60.0-67.0]) improved to 79.0 [70.0-83.0] at 5mg and 4.5 hours post-dose (5mg-4.5H), and worsened to 57.5 [51.0-64.0] at 20mg-1.5H. Baseline Box and Block Test (Modified) median scores (48.0 [47.0-53.0]) worsened to 43.0 [35.0-45.0] at 20mg-1.5H. Baseline Digit Symbol Substitution Test median scores (73.0 [66.0-77.0]) improved to 87.0 [81.0-90.0] at 10mg-4.5H, and worsened to 62.0 [54.0-86.0] at 20mg-1.5H. Reaction Time Ruler Drop Test scores lacked consistent dose-related changes across participants. Changes in states of consciousness were greatest at 20mg. Participants and physiotherapists correctly guessed the administered dose 53% and 50% of the time, respectively.

**Conclusions:** Movement tasks were feasible during psilocybin dosing up to 15mg. Impairments emerged at 20mg in tasks that combined motor and additional cognitive functions. These findings support the feasibility of performing complex movement tasks during psilocybin dosing and will inform the conduct of trials utilising psilocybin-assisted physical rehabilitation in neuropsychiatric disorders.

**Trial Registration:** Australian New Zealand Clinical Trials Registry: ACTRN12621000560897

Date registered: 12 May 2021

URL: https://www.anzctr.org.au/Trial/Registration/TrialReview.aspx?id=381526&isReview=true

**Key Findings:**

- There is growing interest for psychedelic-assisted physical therapy in neuropsychiatric disorders of motor dysfunction, however, the impact of psychedelics on motor function remains unclear.
- This study investigated the feasibility, safety, and impact on motor function of administering movement tasks following low-to-moderate doses of psilocybin in healthy volunteers.
- These findings support the feasibility of performing complex movement tasks during psilocybin dosing up to 15mg and will inform the conduct of trials utilising psilocybin-assisted physical therapy in neuropsychiatric disorders.

## Introduction

Classic psychedelics, such as psilocybin, have been shown to produce widespread effects on brain function [1]. Subjective effects include acutely altered emotion, perception, and consciousness, alongside enduring changes in beliefs and behaviour [2]. Neurobiological effects range from synaptogenesis and the growth of dendritic arbours and spines in cortical neurons [3] to the modulation of macroscopic hierarchical brain dynamics [4]. Psychedelics have shown encouraging treatment outcomes in studies in major depressive disorder, illness-related anxiety, and substance use disorder [5], and are hypothesised to promote structural and functional neuroplasticity that increases the brain’s sensitivity to therapeutic interventions [1,6].

Despite this recent growth in knowledge, very little is known about the impact of psychedelics on motor function. Motor function is underpinned by a dynamic relationship between sensory information and intact higher cognitive functions that support the conceptualisation, planning, and conscious experience of movement [7]. Serotonin-2A receptors, through which psychedelics exert their primary effects [8], are expressed in brain regions associated with motor function, including primary sensory and motor cortices [9] and, at lower levels, the basal ganglia and cerebellum [10,11]. Given this neuroanatomical basis, alongside known changes in sensory perception and macroscopic brain connectivity in networks relevant to motor control following psychedelic administration [12], it is likely psychedelics also impact motor function. However, reports of high-dose psychedelic use in naturalistic settings include preferences for stillness during the acute effects [13], and participants are often instructed to lie down in clinical trials [14], thereby restricting our understanding of motor effects. A previous study assessed motor tasks as part of a wider battery of neurocognitive tests during the acute effects of psilocybin, demonstrating dose-dependent impairments in psychomotor function at 20mg/70kg and 30mg/70kg [15]. However, only two tasks assessing gross motor performance were assessed in this study, and the objective was not to identify a dose at which participants could meaningfully engage in a range of movement tasks during the acute drug effects.

The ability to perform therapeutic movement tasks following psychedelic administration is clinically relevant as psychedelics have been proposed as a treatment for neuropsychiatric disorders associated with motor dysfunction, such as functional neurological disorder (FND), stroke, and acquired brain injury [16–21]. Building upon the growing evidence and recommendations for physical rehabilitation for these disorders [22,23], psychedelic augmentation may facilitate a window of neuroplasticity for optimising therapeutic gains. One treatment model proposes engaging patients in movement retraining during the psychoactive drug effects to take advantage of an early, critical learning period, thought to arise via acute changes in brain dynamics and neuroplasticity following psychedelic administration [4,6,24,25]. However, whether it is feasible to administer a range of movement tasks as required by these therapeutic regimens during the acute psychedelic effects is unknown, and, if feasible, whether a ‘threshold’ dose exists at which individuals can no longer meaningfully engage in this intervention.

Given this theoretical rationale and existing evidence gaps, we investigated the impact of several psilocybin doses on a broad range of motor functions in healthy participants. We hypothesised that a maximum dose exists beyond which participants may be unable to complete movement tasks. Therefore, low-to-moderate doses of psilocybin, below a conventional treatment dose of 25mg, were selected. Doses were administered in a randomised order, with participants, physiotherapists, and statisticians blinded to the order of dosing.

The primary aims were to assess the feasibility and safety of administering a series of movement tasks to healthy volunteers during the acute effects of psilocybin and to identify the maximum dose at which all participants could complete these tasks. Exploratory aims included assessing the impact of low-to-moderate doses of psilocybin on specific domains of motor function, changes in states of consciousness, and blinding efficacy.

## Methods

### Study Design

This is a phase 1, triple-blind, randomised, dose-finding pilot study investigating the effects of low-to-moderate doses of psilocybin on motor function. The study was conducted at Austin Health, a public, tertiary teaching hospital in Melbourne, Australia. Ethical approval was granted by the Austin Health Human Research Ethics Committee. The trial was registered on the Australian New Zealand Clinical Trials Registry (ANZCTR), ACTRN12621000560897. The study protocol was previously published, outlining full details of the study design [26].

### Participants

#### Inclusion criteria

- Adults aged 18 to 65 years.
- Volunteered for the study.
- Capacity to provide informed consent.

#### Exclusion criteria

##### Medical Exclusion Criteria

- Cardiovascular conditions: poorly-controlled hypertension, angina, ischemic heart disease, a clinically significant electrocardiogram abnormality, transient ischemic attack, stroke, or peripheral or pulmonary vascular disease.
- A diagnosis of epilepsy or previous seizures.
- A diagnosis of dementia.
- Chronic kidney or liver disease.
- Known conditions putting the participant at risk for hypercalcaemia, Cushing’s syndrome, hypoglycaemia, syndrome of inappropriate antidiuretic hormone secretion, or carcinoid syndrome.
- Insulin-dependent diabetes: if taking oral hypoglycaemic agents, the participant was only excluded if they also had a history of hypoglycaemia.
- Females who were pregnant, nursing, or trying to conceive.
- Use of medications contraindicated with psilocybin, that were inappropriate to cease for the necessary period surrounding the dosing sessions. See section “Contraindicated Medications” for details.
- Enrolled in another clinical trial involving an investigational product.

##### Psychological Exclusion Criteria

- Current or previous psychotic disorder, including schizophrenia, schizoaffective disorder, schizophreniform disorder, brief psychotic disorder, delusional disorder, schizotypal personality disorder, substance/medication-induced psychotic disorder, or psychotic disorder due to another medical condition.
- Current or previous bipolar I or II disorder.
- First-degree relative with a psychotic or bipolar disorder.
- A history of attempted suicide or mania.
- Current or previous substance use disorder (excluding caffeine and nicotine).
- Previous regular use, or current use, of psychedelic agents.
- Other psychiatric conditions deemed by research staff to be incompatible with safe exposure to psilocybin.

##### Contraindicated medications

- Opioids within 12 hours of psilocybin dosing.
- Antidepressants.
- Potent enzyme inducers or inhibitors.
- Drugs with a narrow therapeutic index within 12 hours of psilocybin dosing.
- Nicotine and caffeine within two hours before and six hours following psilocybin dosing.

#### Study Team Eligibility

The trial physiotherapists were registered physiotherapists trained in the movement tasks assessed in this study. The mental health professionals were medical practitioners with experience working in psychiatric settings.

#### Dose Selection, Randomisation, and Masking

Although no formal classification for the strength of psilocybin doses exists, prior studies typically refer to ‘low’ doses as 10mg or less, ‘moderate’ as 10-20mg, and ‘high’ as above 20mg [27–30], informing the dose selection of 5-20mg as low-to-moderate doses in this study.

The study initially proposed a 3-dose, 3-period cross-over Williams design. The three doses to be examined were 5mg, 10mg, and 15mg of psilocybin, and twelve participants were to be randomly allocated to one of six dosing sequences determined by the Williams design. However, after the first three participants successfully completed the movement tasks at all doses, the dose levels were raised to increase the likelihood of identifying a potential threshold dose at which participants could no longer complete the movement tasks. The protocol was amended, maintaining the same dose range for the first six participants (block one), and changing the dose range to 10mg, 15mg, and 20mg for the last six participants (block two). In doing so, the dose sequences no longer followed a Williams design across all participants.

After the study physician confirmed the participant’s eligibility, the order of each dose was randomised for each participant via a Latin square design within each block. The Research Electronic Data Capture (REDCap) [31–34] randomisation module was used to allocate each sequence, actioned by the study coordinator who was independent of administering the interventions or assessments.

The statisticians, trial physiotherapists assessing the movement tasks, and participants were blinded to the allocation sequence. To maintain blinding, participants were given the same number of capsules for each of their three sessions, comprising a mixture of psilocybin 5mg with or without placebo capsules, contingent on their allocation. The study coordinator, mental health professionals, and study physicians were not blinded, as they arranged the psilocybin prescription and its administration.

#### Procedures

Inquiries were accepted from the community via an advertisement distributed in the Austin Health staff newsletter and the ANZCTR trial listing. Interested participants were provided with the participant information sheet and consent form and invited to a screening visit with a study physician to undertake informed consent, medical and psychiatric screening, and baseline assessments of a series of movement tasks.

Eligible participants then undertook a preparation visit with the study mental health professional to build rapport and discuss information for safe preparation, dosing, and psychological integration, consistent with existing guidelines for safety in psychedelic research [35]. The participant’s order of dosing was then randomised, and they were scheduled for their three dosing sessions, with at least one week between doses.

Psilocybin dosing occurred within a dedicated dosing room, providing a comfortable, monitored, clinical setting with access to temperature control, blankets, headphones, and music. During each dosing session, the participant was encouraged to relax as the drug took effect. They completed a series of movement tasks (see Table 1) at three time points during the acute drug effects (1.5 hours, 3 hours, and 4.5 hours post-dose) with the study physiotherapist. The mental health professional provided supervision and support at all other times, including up to at least 5 hours post-dosing and after the acute drug effects subsided. The mental health professional called the participant the day after each dose to assist in psychological integration and monitor safety. Additionally, a face-to-face psychological integration session was conducted one week after their final dose. Figure 1 provides an outline of this study design.

**Table 1:** Primary and Exploratory Outcomes

|  | Domain | Measures | Application and clinical relevance |
| --- | --- | --- | --- |
| **Primary Outcomes** | Movement tasks | de Morton Mobility Index [36] | 15-item unidimensional instrument that assesses mobility from bed-bound to independent mobility, including balance, seating, and gait. Eleven items are dichotomous (scored 0 or 1), and four items have three response options (scored 0, 1, or 2) based on ability and time to completion. It has high reliability (Pearson's $r=0.94$) and high convergent and discriminant validity. It has a raw score range from 0 to 19, with a conversion table to provide a total score from 0 to 100. Higher scores are indicative of better performance. |
|  |  | Functional Movement Exploration | Movement task measure developed by our team, based on the Physio4FMD intervention in motor functional neurological disorder [43], to assess additional movements, including standing up from a chair, weight shifting, balance, and coordination. Items are dichotomous (scored 0 or 1) based on ability, with additional scores for duration where relevant. Seven items are summed to provide a total score (0-7), or a percentage of the total maximum score (0-100%). Higher scores are indicative of better performance. |
|  | Safety | Vital signs and adverse events form | Monitoring vital signs and recording treatment-emergent adverse events in the adverse events form. These are classified based on the type, number, severity, relatedness to the study drug, and whether the event constitutes a serious adverse event or significant safety issue. |
| **Exploratory Outcomes** | Movement tasks | Action Research Arm Test [37] | 19-item instrument assessing upper limb coordination, dexterity, and functioning. It has shown excellent inter-rater and intra-rater reliability and validity. The total score (range 0-57) is defined as the sum of all 19 items, each scored using a 4-point ordinal scale (range 0-3). Higher scores are indicative of better performance. |
|  |  | Box and Block Test [38] – Original and Modified | • Original: movement of blocks from one box to another over 60 seconds, assessing unilateral gross manual dexterity combined with motor speed. It has shown high test-retest validity from normative data in adults without disability.<br>• Modified: places a shield between the participant's vision and their hands, with access to a mirror to view their movements via reflection. This modulates attentional mechanisms of movement – an important feature of motor functional neurological disorder management.<br>The number of blocks moved in 60 seconds provides the score for each test. Higher scores are indicative of better performance. |
|  |  | Digit Symbol Substitution Test [39] | Symbols are written according to numbers presented via a key within 90 seconds. This recruits a range of cognitive operations, including motor speed, attention, and visuoperceptual functions, and has shown good reliability. The number of correct symbols written in 90 seconds (range 0-90) provides the score. Higher scores are indicative of better performance. |
|  |  | Reaction Time Ruler Drop Test [40] | Participants catch a ruler dropped by the examiner from a vertical height. It has shown excellent test-retest and interrater reliabilities and has been shown to be useful in assessing reaction time. Three attempts are provided. The position on the ruler where the participant catches it (in cm, range 0-30cm) in each attempt is recorded. The average across the three attempts is calculated to provide a mean score. Lower scores are indicative of better performance. |
|  | Altered conscious states | 5-Dimensional Altered States of Consciousness [41] | 94-item scale across eleven dimensions. Responses are rated on a visual analog scale (range 0-100), from 'No, not more than usually' to 'Yes, much more than usually'. The average across items is calculated to provide a mean score overall and by dimension. Higher scores are indicative of greater alterations in consciousness. |
|  |  | Ego-Dissolution Inventory [42] | 8-item focused assessment of the experience of ego-dissolution. Each item is rated on a visual analog scale (range 0-100) from 'No, not more than usually', to 'Yes, entirely or completely'. It has shown discriminant and convergent validity and excellent internal consistency. The average across items is calculated to provide a mean score overall, and individual items represent the score for each dimension. Higher scores are indicative of greater ego-dissolution. |
|  | Blinding | Blinding efficacy questionnaire | Participants and physiotherapists independently guess the dose administered at the end of each dosing session, based upon three possible doses contingent on their block assignment. |

**Figure 1.** 1w: 1 week 5DASC: 5-Dimensional Altered States of Consciousness BS: Baseline (pre-dose) CONSORT: Consolidated Standards of Reporting Trials EDI: Ego-Dissolution Inventory H: Hours post-dose *[Figure was created using Canva (Canva,* https://www.canva.com)]

**Figure 1A:**
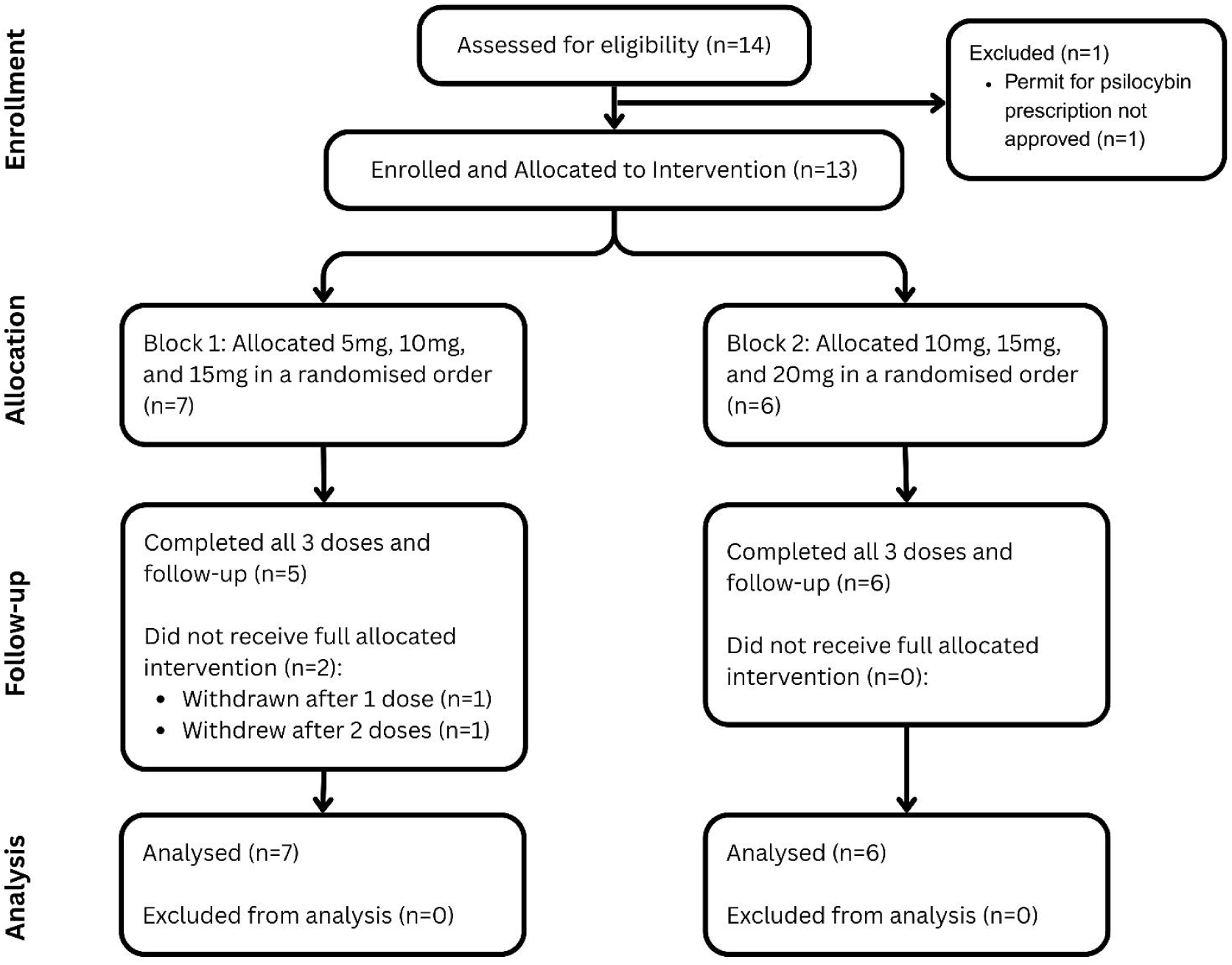
CONSORT Diagram

**Figure 1B:**
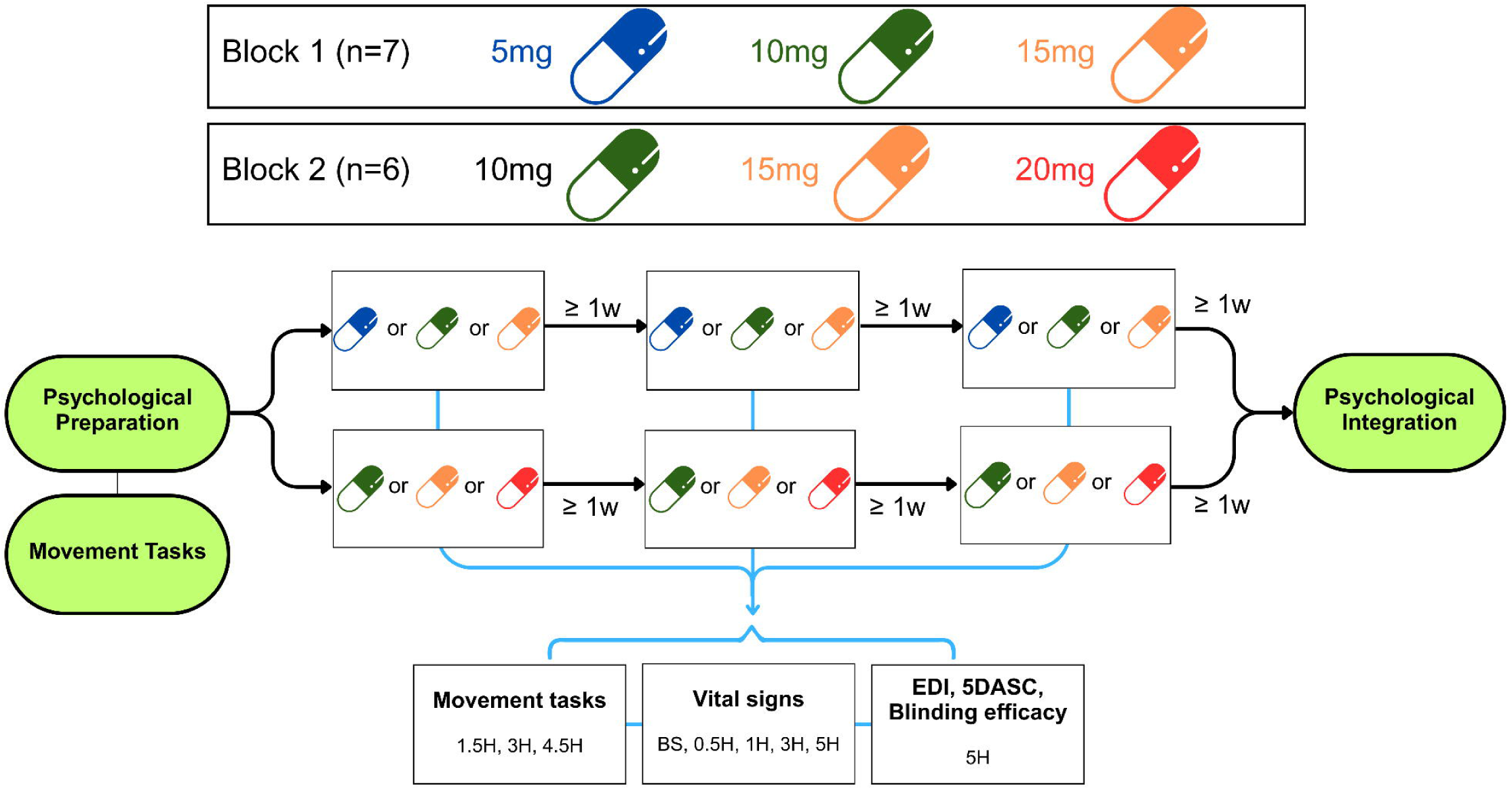
Study Design

#### Outcomes

The primary outcomes were the feasibility of completing the 1) de Morton Mobility Index [36], and 2) Functional Movement Exploration measures of motor function during the acute effects of psilocybin, assessed by task completion and a combined total raw score across both measures (range 0-26).

Safety was assessed by checking vital signs regularly during dosing sessions and recording adverse events at each visit following screening and at any other times arising during the study. Adverse events were classified based on the type, number, severity, relatedness to the study drug, and whether the event constituted a serious adverse event or significant safety issue. A key focus was identifying any adverse events related to administering movement tasks during psilocybin’s acute effects.

Exploratory outcomes included the impact of psilocybin on specific domains of motor function. In addition to the participant’s performance on the de Morton Mobility Index and the Functional Movement Exploration, their performance on the Action Research Arm Test [37], Box and Block Test (Original and Modified versions) [38], Digit Symbol Substitution Test [39], and Reaction Time Ruler Drop Test [40] was assessed. At the end of each dosing session, changes in states of consciousness were measured via the 5-Dimensional Altered States of Consciousness [41] and Ego-Dissolution Inventory [42], and blinding efficacy was assessed by asking the participant and physiotherapist to guess the dose administered. Details of the primary and exploratory outcomes are provided in Table 1.

Additional exploratory outcomes, including verbal fluency, video footage assessing movement quality, resting-state functional magnetic resonance imaging, and qualitative interviews, were also collected and will be reported separately.

### Statistical Methods

The proposed sample size of twelve was not based on statistical power. Rather, this was selected as it was considered sufficient to determine movement task feasibility and performance after each of the three doses of psilocybin.

The analysis included all randomised participants, irrespective of completing their sequence, and all their available non-missing data. No missing data handling techniques were applied. All data were summarised by dose and time point. Outcomes were also summarised by gender. No interim analyses were specified.

For the Functional Movement Exploration, one participant with deafness was unable to conduct the verbal task and, therefore, scored 0 on this item. A sensitivity analysis of the combined total of the de Morton Mobility Index and Functional Movement Exploration score, as well as the Functional Movement Exploration score alone, was conducted, whereby these scores were transformed into a percentage of the maximum total score possible for each participant (range 0-100%), in addition to the total (sum) score. For the Reaction Time Ruler Drop Test, some participants did not grasp the ruler and, therefore, scored a missing value for that attempt. There is no standard convention regarding the scoring of such observations. The final score was averaged over the non-missing scores. A sensitivity analysis was conducted whereby the missing score was set to the length of the ruler (30cm) plus 1cm before averaging across the three attempts.

Post-hoc analyses after unblinding were performed to explore relevant effects and help generate hypotheses for future studies. A linear mixed-effects model was applied with participant specified as a random effect to account for repeated measures. For motor outcomes, a categorical fixed effect representing each dose-time combination was included, with baseline specified as the reference level. This provided an estimate of the mean difference at each dose-time point relative to baseline. For changes in states of consciousness, dose was included as a categorical fixed effect, with 5mg as the reference level, providing estimates of the mean difference in altered states of consciousness for 10mg, 15mg, and 20mg relative to 5mg. In addition to the estimated mean difference, corresponding 95% confidence intervals (CIs) and p-values were provided for all estimates. The full statistical analysis plan is provided in Supplementary Information 1.

## Results

### Participant characteristics

Recruitment occurred over nine months from 18 May 2023 to 7 February 2024. Fourteen participants were screened for eligibility, with one excluded. The remaining thirteen participants were randomised. One participant allocated to block one was withdrawn after one dose due to breaching the study protocol. Accordingly, a seventh participant was enrolled into block one to maintain the plan for six participants to complete the block. The subsequent six participants were assigned to block two. One participant in block one withdrew after two doses due to competing external commitments. Eleven participants completed all three allocated doses. Available, non-missing data for all thirteen participants were included in the analysis (see Fig. 1). The mean age of enrolled participants was 31 years (range 22-39 years), the mean height was 173cm (160-182cm), the mean weight was 71kg (51-94kg), and five (62%) participants were male and eight (38%) were female.

### Primary outcomes

Hereafter, dose-time points are denoted using the format dose-time (for example, 20mg at 3 hours post-dose is written as 20mg-3H).

In terms of the feasibility of completing the de Morton Mobility Index and the Functional Movement Exploration, one participant experienced pronounced subjective drug effects and movement difficulty at 20mg and was unable to begin several movement tasks at 20mg-1.5H and all tasks at 20mg-3H. Specifically, the participant reported reduced perception of their physical boundaries and remained lying on the floor with blankets wrapped around them to maintain their sense of safety and provide peripheral, sensory feedback. Despite repeated checks by the mental health professional, these experiences were not reported as distressing or adverse. This participant’s combined total raw score for these two measures was 16/26 at 20mg-1.5H and 0/26 at 20mg-3H. All other participants who were administered both measures completed all tasks at all assessed dose-time points, achieving combined total raw scores ranging from 24/26 to 26/26 (see Fig. 2A).

**Figure 2.**
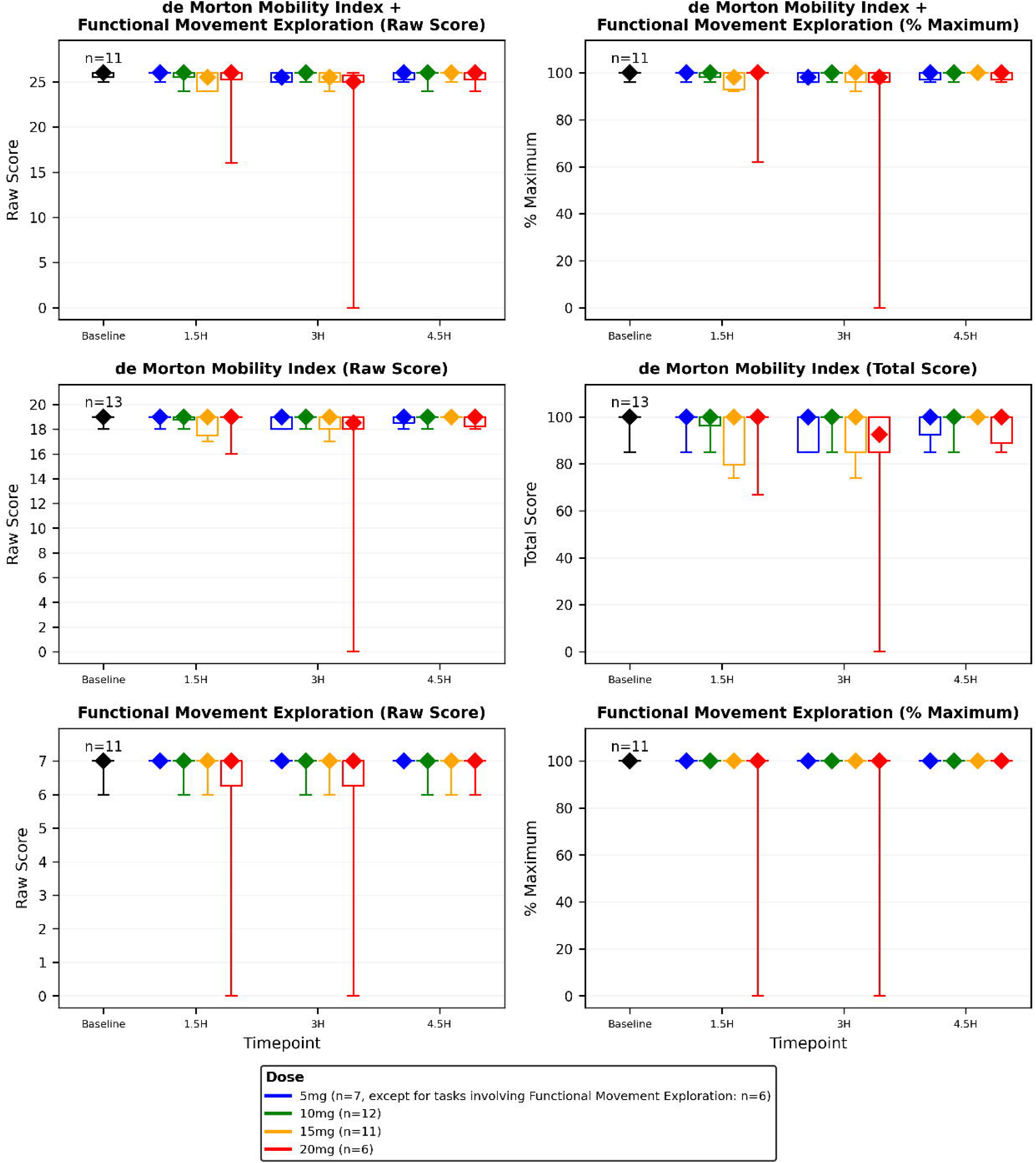
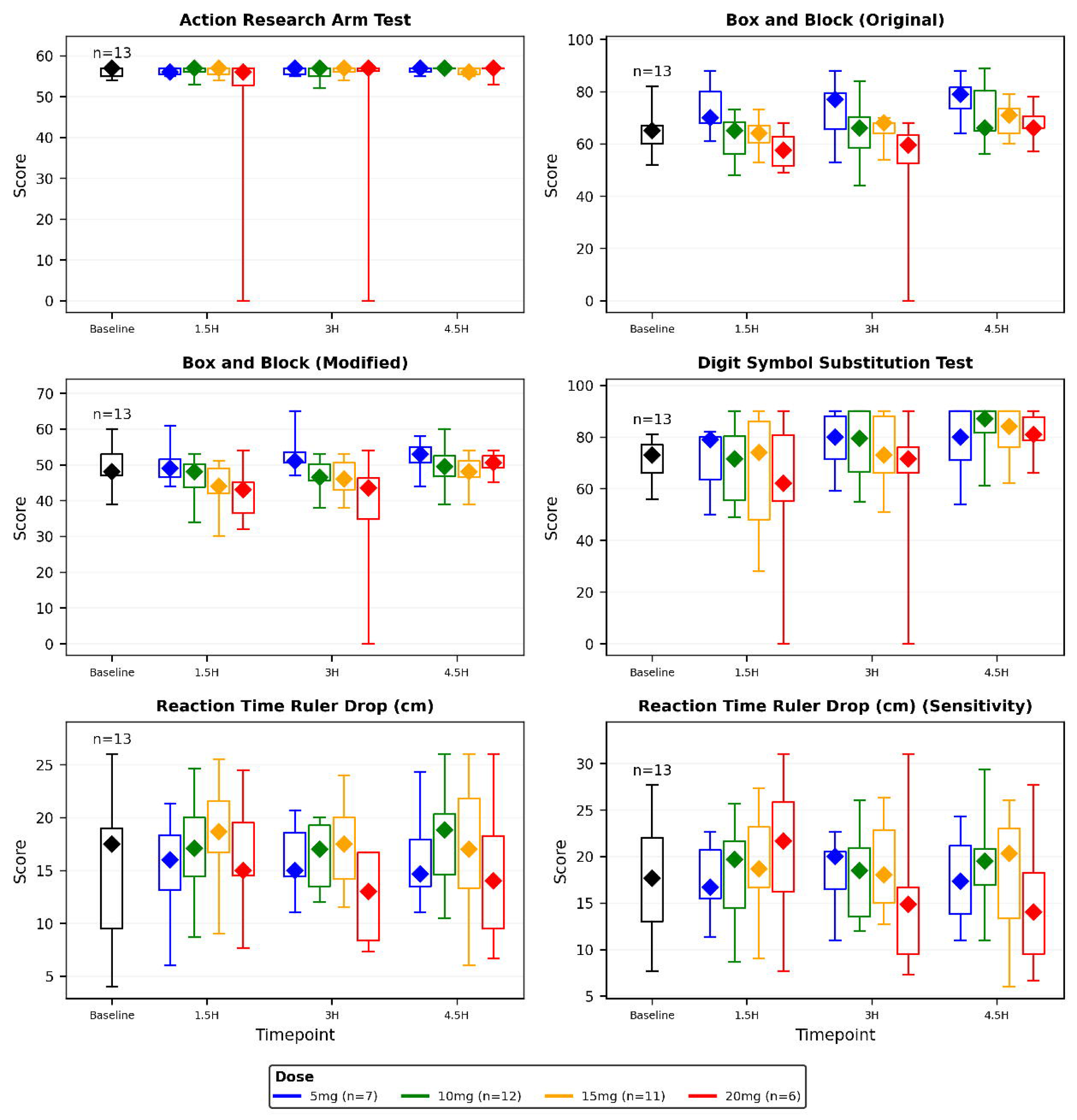
Diamonds: median value Boxes: Interquartile range Whiskers: Full range H: Hours post-dose *[Figure was generated using Python (version 3.13.2), with the following libraries: matplotlib and pandas]*

No adverse events occurred that were related to the administration of movement tasks during the acute drug effects. No serious adverse events (events that result in death, are life-threatening, require hospitalisation or prolong existing hospitalisation, cause persistent or significant disability or incapacity, or involve a congenital anomaly or birth defect) or significant safety issues (adversely affect the safety of participants or materially impact on the continued ethical acceptability or conduct of the trial) occurred throughout the study. A total of 64 adverse events were reported throughout the study from eleven (85%) participants, including 62 (97%) considered related to psilocybin. 6 adverse events (9%) were of nil severity (no interference with everyday activities), 52 (81%) were mild (tolerated by the participant, causing minimal discomfort and not interfering with everyday activities), 5 (8%) were moderate (sufficiently discomforting to interfere with everyday activities), and 1 (2%) was severe (prevents everyday activities). The sole severe adverse event occurred in the evening after the participant returned home following their 15mg dose. The event consisted of nausea and vomiting after their dinner and was associated with a probable brief syncopal episode. A foodborne illness was considered the most likely explanation for this event, given the timeframe; however, contributions from psilocybin could not be fully excluded, and, therefore, the event was graded as possibly related to psilocybin. All moderate and severe adverse events were associated with the 15mg dose.

39 (61%) adverse events occurred during the dosing session, with the remaining 25 (39%) adverse events occurring post-dosing, typically within 24 hours. Of all adverse events, nausea (with or without vomiting) and headache were the most common, occurring in eight (62%) and seven (54%) participants, respectively. Median values for blood pressure and heart rate increased from baseline during the acute effects of dosing, with no recordings warranting acute medical intervention. No rescue medications were required throughout the study. Further details regarding safety data are provided in Table 2 and Supplementary Information 2.

**Table 2:** Adverse Events

|  | **5mg** | **10mg** | **15mg** | **20mg** | **Total** |
| --- | --- | --- | --- | --- | --- |
|  | N=7 | N=12 | N=11 | N=6 | N=13 |
| **Table 2A: Number of participants with adverse events during enrolment** |  |  |  |  |  |
| At least any adverse event | 3 (43%) | 8 (67%) | 8 (73%) | 4 (67%) | 11 (85%) |
| At least one related* adverse event | 3 (43%) | 8 (67%) | 8 (73%) | 4 (67%) | 11 (85%) |
| At least one severe adverse event | 0 (0%) | 0 (0%) | 1 (9%) | 0 (0%) | 1 (8%) |
| At least one adverse event during dosing | 3 (43%) | 8 (67%) | 8 (73%) | 3 (50%) | 11 (85%) |
| At least one related* adverse event during dosing | 3 (43%) | 8 (67%) | 8 (73%) | 3 (50%) | 11 (85%) |
| At least one severe adverse event during dosing | 0 (0%) | 0 (0%) | 0 (0%) | 0 (0%) | 0 (0%) |
| At least one adverse event post-dosing | 2 (29%) | 3 (25%) | 5 (45%) | 3 (50%) | 7 (54%) |
| At least one related* adverse event post-dosing | 2 (29%) | 3 (25%) | 5 (45%) | 3 (50%) | 7 (54%) |
| At least one severe adverse event post-dosing | 0 (0%) | 0 (0%) | 1 (9%) | 0 (0%) | 1 (8%) |
| **Table 2B: Number and type of adverse events during enrolment** |  |  |  |  |  |
| **Number of adverse events** | 10 (100%) | 19 (100%) | 24 (100%) | 11 (100%) | 64 (100%) |
| **Name of adverse event** |  |  |  |  |  |
| Anxiety | 0 (0%) | 0 (0%) | 0 (0%) | 2 (18%) | 2 (3%) |
| Body ache | 0 (0%) | 1 (5%) | 0 (0%) | 1 (9%) | 2 (3%) |
| Dizziness | 0 (0%) | 1 (5%) | 2 (8%) | 1 (9%) | 4 (6%) |
| Drowsiness or sedation | 1 (10%) | 2 (11%) | 2 (8%) | 0 (0%) | 5 (8%) |
| Dry mouth | 0 (0%) | 0 (0%) | 1 (4%) | 0 (0%) | 1 (2%) |
| Feeling cold inside | 0 (0%) | 0 (0%) | 1 (4%) | 0 (0%) | 1 (2%) |
| Feeling flushed in the face | 0 (0%) | 0 (0%) | 1 (4%) | 0 (0%) | 1 (2%) |
| Feeling run down | 1 (10%) | 0 (0%) | 0 (0%) | 0 (0%) | 1 (2%) |
| Hand tremor | 0 (0%) | 1 (5%) | 0 (0%) | 0 (0%) | 1 (2%) |
| Headache | 1 (10%) | 2 (11%) | 3 (12%) | 2 (18%) | 8 (12%) |
| Hot and cold | 0 (0%) | 0 (0%) | 1 (4%) | 0 (0%) | 1 (2%) |
| Increased urinary frequency | 1 (10%) | 1 (5%) | 1 (4%) | 0 (0%) | 3 (5%) |
| Jaw stiffness | 0 (0%) | 1 (5%) | 0 (0%) | 0 (0%) | 1 (2%) |
| Lightheadedness | 0 (0%) | 1 (5%) | 1 (4%) | 0 (0%) | 2 (3%) |
| Muscle soreness | 1 (10%) | 0 (0%) | 0 (0%) | 0 (0%) | 1 (2%) |
| Nausea with or without vomiting | 3 (30%) | 4 (21%) | 5 (21%) | 1 (9%) | 13 (20%) |
| Restlessness | 0 (0%) | 0 (0%) | 1 (4%) | 0 (0%) | 1 (2%) |
| Rhinorrhoea and lacrimation | 0 (0%) | 1 (5%) | 1 (4%) | 1 (9%) | 3 (5%) |
| Sound sensitivity | 0 (0%) | 0 (0%) | 0 (0%) | 1 (9%) | 1 (2%) |
| Tiredness | 1 (10%) | 1 (5%) | 1 (4%) | 1 (9%) | 4 (6%) |
| Warmth | 0 (0%) | 0 (0%) | 0 (0%) | 1 (9%) | 1 (2%) |
| Weakness | 0 (0%) | 2 (11%) | 2 (8%) | 0 (0%) | 4 (6%) |
| Yawning | 1 (10%) | 1 (5%) | 1 (4%) | 0 (0%) | 3 (5%) |
| **Severity of adverse event** |  |  |  |  |  |
| Nil | 3 (30%) | 2 (11%) | 0 (0%) | 1 (9%) | 6 (9%) |
| Mild | 7 (70%) | 17 (89%) | 18 (75%) | 10 (91%) | 52 (81%) |
| Moderate | 0 (0%) | 0 (0%) | 5 (21%) | 0 (0%) | 5 (8%) |
| Severe | 0 (0%) | 0 (0%) | 1 (4%) | 0 (0%) | 1 (2%) |
| **Serious adverse event** |  |  |  |  |  |
| Any | 0 (0%) | 0 (0%) | 0 (0%) | 0 (0%) | 0 (0%) |
| **Relatedness of adverse event to psilocybin** |  |  |  |  |  |
| not assessable | 1 (10%) | 1 (5%) | 0 (0%) | 0 (0%) | 2 (3%) |
| possibly related | 3 (30%) | 11 (58%) | 15 (62%) | 7 (64%) | 36 (56%) |
| probably related | 6 (60%) | 7 (37%) | 8 (33%) | 2 (18%) | 23 (36%) |
| definitely related | 0 (0%) | 0 (0%) | 1 (4%) | 2 (18%) | 3 (5%) |

\*related: Relatedness to psilocybin Table 2A: Values refer to the number (%) of participants. Table 2B Values refer to the number (%) of adverse events.
\*related: Relatedness to psilocybin Table 2A: Values refer to the number (%) of participants.

### Exploratory outcomes

Beyond assessments of feasibility and combined scores, analyses of individual movement tasks revealed dose-dependent effects within time points, ranging from improvements at lower doses for some measures to impairments at the highest dose, supported by post-hoc statistical analyses. The participant who was unable to engage in several movement tasks at 20mg scored 67/100, 0/100%, and 0/57 for the de Morton Mobility Index (total score), Functional Movement Exploration (% maximum), and Action Research Arm Test, respectively, at 20mg-1.5H; and 0/100, 0/100%, and 0/57 for these measures at 20mg-3H. All other participants scored between 74/100 and 100/100 for the de Morton Mobility Index, their maximum possible score (100%) for the Functional Movement Exploration, and between 52/57 and 57/57 for the Action Research Arm Test at all dose-time points. For the Reaction Time Ruler Drop Test, median values lacked consistent dose-related changes across participants, with similar findings observed in the corresponding sensitivity analysis.

Tests combining motor speed with dexterity or additional cognitive functions demonstrated greater impairments at 20mg (peak effects), and improvements at 5mg and/or 10mg (post-peak effects). Median scores [interquartile ranges] for the Box and Block Test (Original) increased (improved) from baseline (65.0 [60.0-67.0]) to 79.0 [70.0-83.0] at 5mg-4.5H, and decreased (worsened) to 57.5 [51.0-64.0] at 20mg-1.5H. Box and Block Test (Modified) median scores decreased (worsened) from baseline (48.0 [47.0-53.0]) to 43.0 [35.0-45.0] at 20mg-1.5H. Digit Symbol Substitution Test median scores increased (improved) from baseline (73.0 [66.0-77.0]) to 87.0 [81.0-90.0] at 10mg-4.5H, and decreased (worsened) to 62.0 [54.0-86.0] at 20mg-1.5H. For these three measures, median values were improved at all time points at 5mg and worsened during peak effects at 20mg compared to baseline.

Figure 2A presents descriptive statistics for the de Morton Mobility Index and Functional Movement Exploration, including combined scores, individual raw and scaled scores, and corresponding sensitivity analyses. Figure 2B presents the descriptive statistics for the remaining movement tasks, including the original and sensitivity analyses for the Reaction Time Ruler Drop Test.

Plots of individual scores are provided in Supplementary Information 3. Subgroup analyses did not identify consistent, clinically meaningful differences by gender; descriptive statistics by gender are provided in Supplementary Information 4.

Measures of changes in states of consciousness via the 5-Dimensional Altered States of Consciousness and Ego-Dissolution Inventory revealed greater differences in several domains at 20mg compared to other doses (See Fig. 3). For the 5-Dimensional Altered States of Consciousness, this included greater changes in insightfulness, disembodiment, complex imagery, and elemental imagery. For the Ego-Dissolution Inventory, this included higher scores for items describing disintegration, dissolution, and a loss of one’s self or ego.

**Figure 3.**
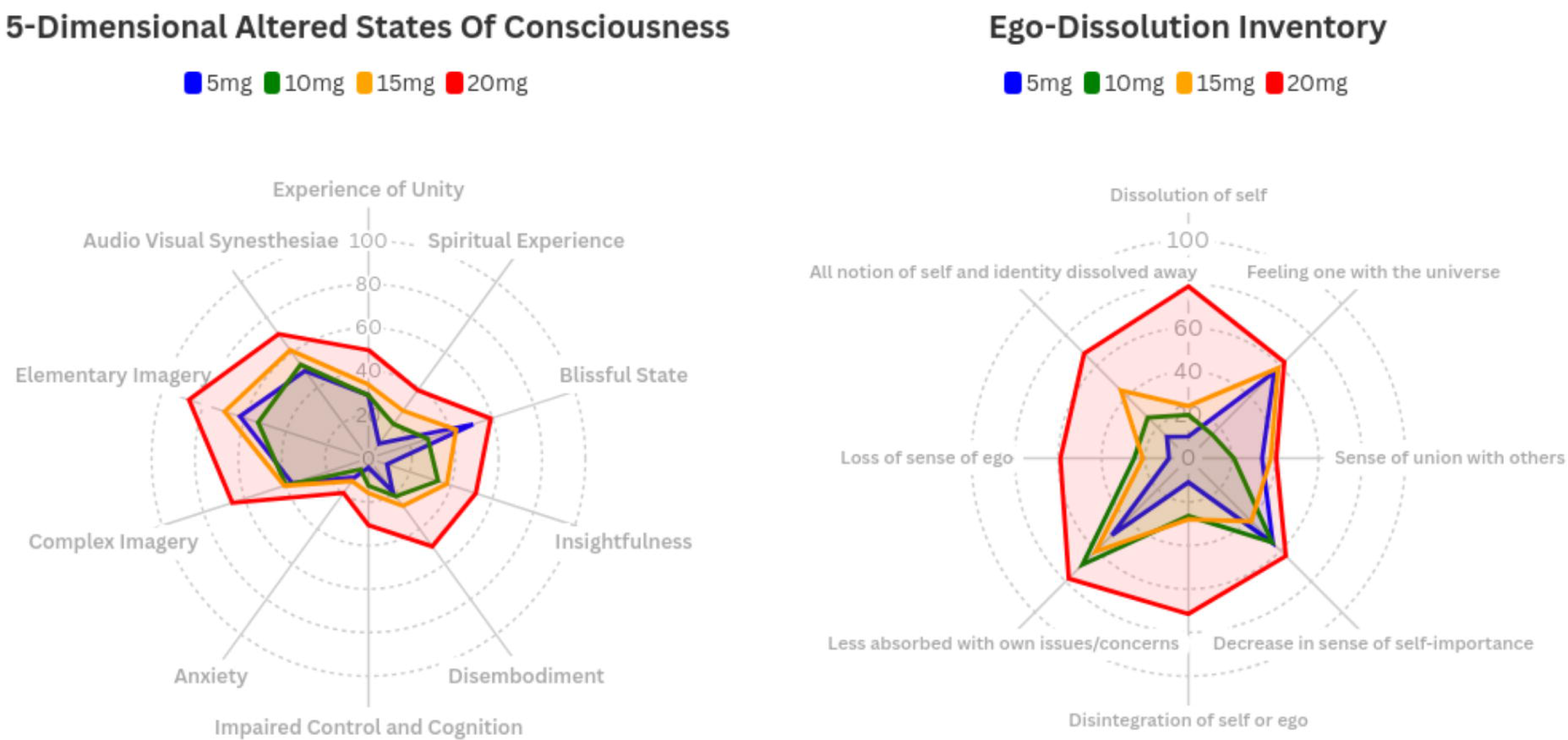
[Plots were generated using Flourish (https://flourish.studio), and final figure layout and annotation were completed in Canva (https://www.canva.com)]

Regarding blinding efficacy, participants and physiotherapists guessed the dose administered correctly 53% and 50% of the time, respectively.

### Post-Hoc Analyses

Due to the ceiling effect and very little variation in the de Morton Mobility Index, Functional Movement Exploration, and Action Research Arm Test scores, the linear mixed-effects model provided a poor fit for these outcomes; therefore, the results of these post-hoc statistical analyses are not reported here.

Exploratory statistical testing for the Box and Block Tests demonstrated statistically significant (p-value less than the nominal level of 0.05) impairments at 20mg-3H (Box and Block Test Original: mean difference (MD) = -11.0, 95% CI = [-20.0, -2.9], p = 0.009; Box and Block Test Modified: MD = - 11.0, 95% CI = [-18.0, -5.3], p < 0.001). In addition, for the Box and Block Test Original, there were statistically significant improvements at 5mg-4.5H (MD = 11.0, 95% CI = [2.6, 18.0], p = 0.010) and 10mg-4.5H (MD = 7.4, 95% CI = [0.8, 14.0], p = 0.029). The degree of impairment (11 fewer blocks) and improvement (7-11 more blocks) is greater than the proposed, minimal clinically important differences of up to 7 blocks reported in clinical settings [44,45]. For the Digit Symbol Substitution Test, statistically significant impairments were seen at 20mg-1.5H (MD = -16.0, 95% CI = [-27.0, -4.9], p = 0.005) and 20mg-3H (MD = -12.0, 95% CI = [-23.0, -1.4], p = 0.028), and improvement at 10mg-4.5H (MD = 10.0, 95% CI = [1.3, 19.0], p = 0.025), which constituted greater changes in both directions than minimal clinically important differences of between 3-6 symbols reported in clinical settings. These minimal clinically important differences should be interpreted cautiously, as they were tested in other clinical cohorts, and their applicability in this context is uncertain. Moreover, the participant who was unable to begin all tasks at 20mg-3H due to profound movement impairment scored 0 for these three measures, likely contributing substantially to these effects (see plots of individual scores provided in Supplementary Information 3). For the Reaction Time Ruler Drop Test, no statistically significant changes were observed. For this analysis, the original scoring method was applied, whereby attempts when participants did not grasp the ruler were recorded as missing values. Estimates of the mean change from baseline for these movement tasks derived from statistical modelling are plotted in Figure 4.

**Figure 4.**
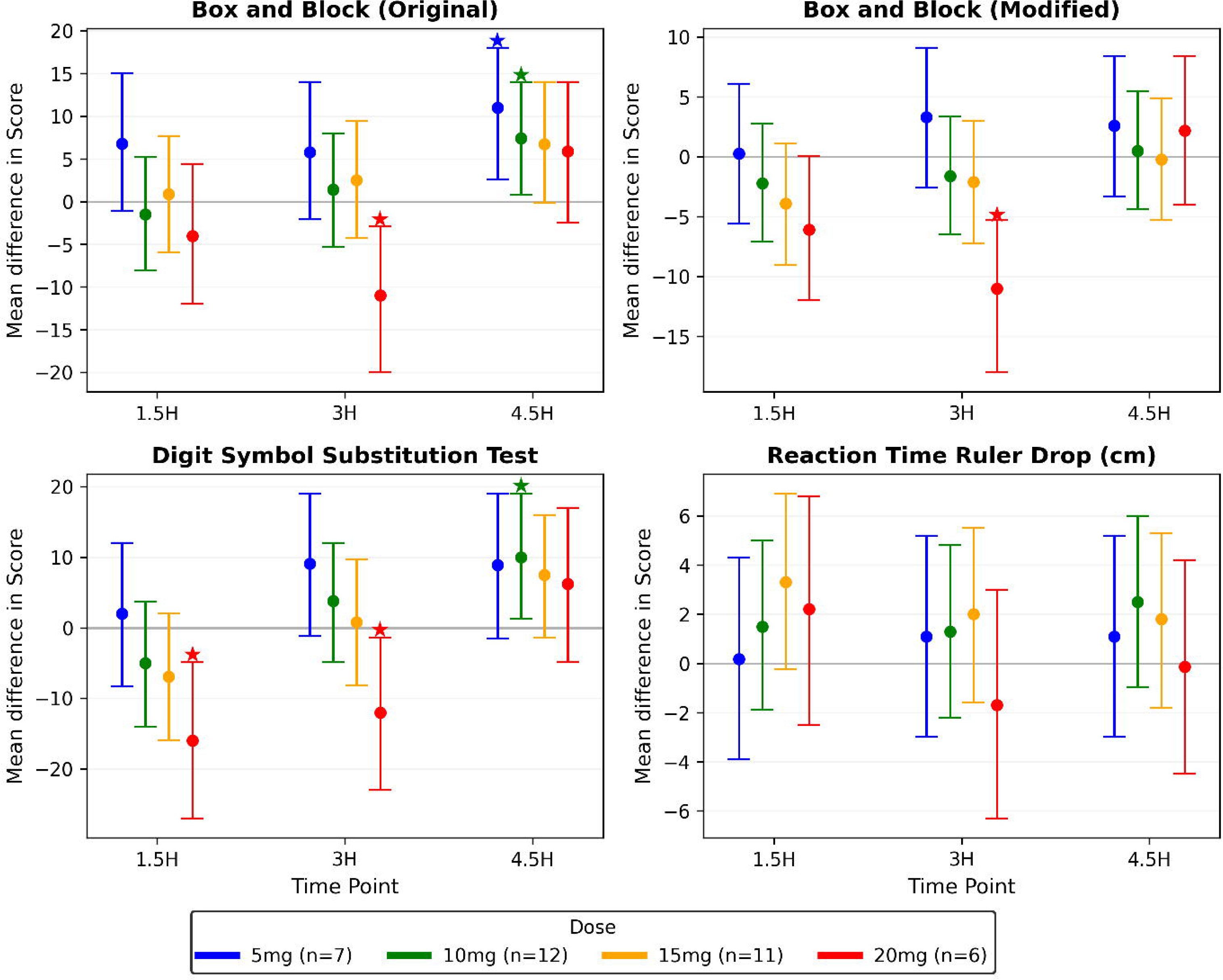
Points represent the estimated mean difference from baseline for each dose and timepoint compared to the reference category (no difference is denoted by the grey line), with vertical lines indicating 95% confidence intervals. Asterisks (*) indicate statistically significant effects (p-value < 0.05). H: Hours post-dose *[Figure was generated using Python (version 3.13.2), with the following libraries: matplotlib and pandas]*

For the 5-Dimensional Altered States of Consciousness, statistically significant increases were seen at 15mg and 20mg, relative to 5mg (15mg: MD = 9.9, 95% CI = [1.7, 18.0], p = 0.020; 20mg: MD = 14.0, 95% CI = [4.5, 24.0], p = 0.007). Subscales showing statistically significant increases included experience of unity (20mg), audio-visual synaesthesia (15mg and 20mg), and changed meaning of percepts (20mg). For the Ego-Dissolution Inventory, statistically significant increases in the mean total score were seen at 15mg and 20mg, relative to 5mg (15mg: MD = 17.0, 95% CI = [2.1, 33.0], p = 0.028; 20mg: MD = 34.0, 95% CI = [16.0, 53.0], p < 0.001). Subscales showing statistically significant increases included dissolution of self (20mg), disintegration of self or ego (20mg), less absorbed by own issues or concerns (20mg), loss of sense of ego (10mg and 20mg), and notion of self and identity having dissolved away (15mg and 20mg). While no minimal clinically important differences were identified for these measures, the magnitude of mean differences at 20mg compared to 5mg (≥14 points on a 0-100 scale) likely reflects meaningful effects at this dose. Further details for these statistical tests are provided in Supplementary Information 5.

## Discussion

This is the first study to systematically evaluate the impact of psilocybin on motor function and the feasibility of performing movement tasks during acute psychedelic drug effects. We found that it was feasible to administer a series of movement tasks to healthy participants during the acute drug effects up to a dose of 15mg. Beyond this, greater impairments were seen during the peak drug effects, especially in tasks combining multiple motor and cognitive functions, accompanied by greater changes in states of consciousness. In contrast, possible improvements in motor function were observed at lower doses after the peak drug effects subsided.

No adverse events occurred in this study that were directly related to movement task administration, and there were no serious adverse events. Most adverse events were mild and transient, with nausea and headache the most common, occurring in eight (62%) and seven (54%) participants, respectively. These findings are comparable to other studies of psilocybin in healthy and clinical populations that demonstrate short-term tolerability [46–51]. Regarding feasibility, all participants could complete movement tasks between 5mg and 15mg (inclusive). However, one participant was unable to begin several movement tasks during the peak effects at 20mg, underscoring the potential for profound immobility at this dose. This may be consistent with individual preferences for stillness in high-dose recreational settings [13], and clinical instructions for participants to remain still during clinical trials utilising conventional treatment doses of approximately 25mg [14], perhaps in anticipation of these movement difficulties at these doses.

In terms of specific motor functions, the de Morton Mobility Index and Functional Movement Exploration assess several gross motor movements, including balance, gait, and sit-to-stand movements [36], while the Action Research Arm Test measures upper limb coordination and dexterity [37]. For these gross and fine motor functions, besides one participant who was unable to engage in movement tasks during the peak drug effects at 20mg, all other participants showed minimal change across doses. Consistent, dose-related changes were also not observed across the sample in reaction time, as assessed by the Reaction Time Ruler Drop Test.

However, impairments were observed in tasks that combine multiple motor and/or additional cognitive functions. The Box and Block Tests [38] (Original and Modified versions) combine dexterity with motor speed, while the Digit Symbol Substitution Test combines motor speed with attention and visuo-perceptual functions. These impairments arising at 20mg during the peak drug effects may signal a potential threshold at which psilocybin impairs these motor functions. In contrast, improvements were seen at lower doses for both the Box and Block Test (Original) and the Digit Symbol Substitution Test, especially after the peak effects subsided. Although contributions via practice effects developing during each dosing session are possible, these improvements, albeit modest, are noteworthy in an already healthy cohort and warrant further inquiry.

Synthesising these findings, the lack of impairments in the de Morton Mobility Index, Functional Movement Exploration, Action Research Arm Test, and Reaction Time Ruler Drop Test may suggest sufficient capacity is retained for these relatively isolated motor tasks. However, the stronger effects of psilocybin at 20mg may diminish the ability to recruit and coordinate multiple motor and associated cognitive functions simultaneously, overwhelming the brain’s capacity to execute more complex tasks incorporated within the Box and Block Tests and Digit Symbol Substitution Test.

These impairments may complement the substantially greater changes in states of consciousness and ego-dissolution at higher doses, reflecting widespread changes in higher-order cortical functions and their potential association with these complex motor functions. While the qualitative analysis of participant interviews is being prepared separately from this paper, preliminary findings reflect dose-dependent effects that support these objective measures of motor function. These include reports of improvements at lower doses and difficulties at higher doses. Notably, these subjective impairments were influenced by perceived alterations in associated cognitive functions and conscious states, such as diminished attentional capacity and mystical experiences.

These motor function results align with the findings from the only other study we identified that incorporated select movement tasks (within a broader battery of neurocognitive measures) during acute psychedelic effects by Barrett and colleagues [15]. In their study, one task assessing fine motor accuracy and response time, two gross motor tasks, and the Digit Symbol Substitution Test were administered in 20 healthy volunteers during the acute effects of several drug conditions (psilocybin at three doses: 10mg/70kg, 20mg/70kg, and 30mg/70kg; and dextromethorphan 400mg/70kg) compared to placebo. Statistically significant impairments were observed during peak effects (2 hours post-dose) at psilocybin 20mg/70kg and 30mg/70kg compared to placebo for response time, both gross motor tasks, and the Digit Symbol Substitution Test, but not for fine motor accuracy. Compared to our study, these measures comprised a limited set of motor function assessments, possibly reducing the ability to detect subtle changes. Nevertheless, these findings support our observations of more marked impairments emerging only once the psilocybin dose reaches 20mg.

Moreover, these effects of psilocybin on motor function may differ in quality from those of other psychoactive substances. In the study by Barrett and colleagues, dextromethorphan 400mg/70kg often produced greater impairments than most psilocybin doses in gross motor tasks [15]. Similarly, alcohol intoxication is known to more directly impair multiple domains of motor control, including gait, coordination, balance, speech, and psychomotor speed [52]. Drawing upon their known pharmacological actions, dextromethorphan and alcohol inhibit N-methyl-D-aspartate receptor function [53,54], with alcohol additionally potentiating γ-aminobutyric acid type-A receptors [55], converging upon disrupted glutaminergic neurotransmission throughout the central nervous system, including areas directly involved in movement execution and motor control, such as the basal ganglia, thalamus, brainstem, and cerebellum [53,55,56]. In contrast, psilocybin’s action on serotonin-2A receptors, which are more densely expressed in higher-order cortical areas, may reflect its differential effects on associative cognitive functions related to movement, such as planning, motivation, and intent.

This study’s findings may also be consistent with neuroimaging evidence of acutely altered brain dynamics following psychedelic administration, and present intriguing opportunities that could be leveraged in future studies [6,57–60]. At a local level, psychedelics give rise to acute entropic effects on spontaneous cortical activity, enabling neural dynamics to deviate from typical patterns of activity. These findings are accompanied by widespread desynchronisation in brain function, typically characterised by reduced within-network and increased between-network functional connectivity across cortical, resting-state networks. In the context of motor function, substantial deviations during peak effects at stronger doses may therefore compromise the usually coordinated activity involving motor and associated cognitive areas involved in these more complex movements. Conversely, at lower doses, this may foster the revision of previously entrenched pathways and the emergence of novel approaches to movement. These improvements may materialise as the peak effects subside, the acutely entropic state passes, and neural dynamics restabilise with renewed properties.

This is of clinical relevance given the hypothesised role for psychedelics combined with movement retraining for neuropsychiatric disorders associated with motor dysfunction [16]. While grounded in a theoretical basis to take advantage of these acute changes in the brain dynamics following psychedelic administration [4,6,24], psilocybin 20mg may represent a threshold dose whereby some individuals are no longer able to engage meaningfully in physical therapy. This is important as disorders that may benefit from this approach, such as motor FND, acquired brain injury, Parkinson’s disease, and stroke [16–18,20,21,25], inherently require physical rehabilitation that simultaneously recruits multiple brain functions based upon their underlying pathophysiology. Motor FND, for example, is characterised by motor symptoms, such as abnormal gait, weakness, or tremor, that are considered incompatible with other neurological disorders [61], and expert consensus recommends psychologically-informed physiotherapy as part of a multidisciplinary approach [23]. The physiotherapy approach emphasises associated intellectual and cognitive functions to augment movement retraining, such as promoting insight via psychoeducation and utilising attentional diversion to elicit automatic movements [43]. Therefore, if adopting physical rehabilitation during the acute psychedelic effects, the dose selection will need to balance promoting neuroplastic effects while retaining sufficient brain capacity for meaningful engagement in these biopsychosocial interventions.

Although significant carryover effects between each dose were not anticipated, randomisation and blinding to the order of dosing assisted in balancing any such effects arising across doses. Furthermore, this approach mitigated expectancy effects, evidenced by both participants and physiotherapists guessing the dose correctly only about half the time. Given the public interest in psychedelic research and the influence of expectations on research outcomes [62], these findings reflect the strengths of the randomisation in this study and the potential utility of this approach to mitigate expectancy and placebo confounds in future psychedelic studies.

This study is limited by the small sample size and the ceiling effect of several included movement tasks, which may have constrained the ability to detect nuanced changes across dose-time points. Although impressive blinding efficacy was observed, participants and physiotherapists guessed the dose correctly at a rate greater than chance alone. Moreover, no placebo was offered; therefore, a degree of expectancy was likely and could not be fully excluded.

In conclusion, this investigation supports the feasibility of administering movement tasks during the acute effects of psilocybin to healthy participants up to a dose of 15mg. These findings provide the foundation for future hypothesis-driven studies of psilocybin on specific domains of motor function, and for testing psilocybin combined with movement retraining in clinical populations.

## Statements and Declarations

### Funding

This work was supported by the Medical Research Future Fund (R.K., O.C., S.B., G.N., D.B., and A.B., grant number MRF2012410); the Wellcome Trust (M.B., grant number 227515/Z/23/Z), and the RANZCP Foundation, the Royal Australian and New Zealand College of Psychiatrists (C.B.).

The Usona Institute provided the study drug. They have not offered or provided payments to the investigators.

### Trial Sponsor

Austin Health, Austin Hospital, 145 Studley Rd, Heidelberg VIC 3084.

### Ethical Approval

Ethical approval for this protocol has been awarded by the Austin Health Human Research Ethics Committee (HREC/57390/Austin-2020).

The authors assert that all procedures contributing to this work comply with the ethical standards of the relevant national and institutional committees on human experimentation and with the Helsinki Declaration of 1964 and its later amendments.

### Consent

Informed consent was obtained from all individual participants for participation in this study and publication of their deidentified data.

### Data Availability

Relevant data has been included as supplementary information. Further details may be provided by the corresponding author upon reasonable request.

### Authors’ contributions

R.K. is the Principal Investigator and the senior researcher. C.B., O.C., G.N., D.B., S.I., S.B., S.Z., Z.A., G.O., D.M., J.R., M.B., A.B., and R.A.K. contributed to the development of the protocol. C.B., R.K., and Z.A. collected the data. C.B., S.B, and S.Z. developed the statistical analysis plan and conducted the analyses. All authors contributed to the interpretation of the findings. C.B. drafted the manuscript. A.B. and R.A.K. provided supervision. All authors approved the final manuscript.

