## Supplementary Information 1 for "A Randomised, Triple-Blind, Dose-Finding Study of the Impact of Psilocybin on Motor Function in Healthy Participants"

### Supplementary Information 2: Safety Data

### Adverse Events

**Table S2.1: Adverse Events**

|  | **5mg** | **10mg** | **15mg** | **20mg** | **Total** |
| --- | --- | --- | --- | --- | --- |

| **Adverse event or adverse reaction during study** |  |  |  |  |  |
| --- | --- | --- | --- | --- | --- |
| adverse event (an event which is unrelated to psilocybin) | 1 (10%) | 1 (5%) | 0 (0%) | 0 (0%) | 2 (3%) |
| adverse reaction (an event which is possibly/probably/definitely related to psilocybin) | 9 (90%) | 18 (95%) | 24 (100%) | 11 (100%) | 62 (97%) |
| **Significant Safety Issue during study** |  |  |  |  |  |
| No | 10 (100%) | 19 (100%) | 24 (100%) | 11 (100%) | 64 (100%) |
| **Adverse event during dosing** | 6 (60%) | 15 (79%) | 13 (54%) | 5 (45%) | 39 (61%) |
| **Name of adverse event during dosing** |  |  |  |  |  |
| Anxiety | 0 (0%) | 0 (0%) | 0 (0%) | 2 (40%) | 2 (5%) |
| Dizziness | 0 (0%) | 1 (7%) | 2 (15%) | 1 (20%) | 4 (10%) |
| Drowsiness or sedation | 0 (0%) | 2 (13%) | 1 (8%) | 0 (0%) | 3 (8%) |
| Dry mouth | 0 (0%) | 0 (0%) | 1 (8%) | 0 (0%) | 1 (3%) |
| Hand tremor | 0 (0%) | 1 (7%) | 0 (0%) | 0 (0%) | 1 (3%) |
| Increased urinary frequency | 1 (17%) | 1 (7%) | 1 (8%) | 0 (0%) | 3 (8%) |
| Jaw stiffness | 0 (0%) | 1 (7%) | 0 (0%) | 0 (0%) | 1 (3%) |
| Lightheadedness | 0 (0%) | 1 (7%) | 0 (0%) | 0 (0%) | 1 (3%) |
| Muscle soreness | 1 (17%) | 0 (0%) | 0 (0%) | 0 (0%) | 1 (3%) |
| Nausea with or without vomiting | 2 (33%) | 4 (27%) | 4 (31%) | 1 (20%) | 11 (28%) |
| Restlessness | 0 (0%) | 0 (0%) | 1 (8%) | 0 (0%) | 1 (3%) |
| Rhinorrhoea and lacrimation | 0 (0%) | 1 (7%) | 1 (8%) | 1 (20%) | 3 (8%) |
| Tiredness | 1 (17%) | 0 (0%) | 0 (0%) | 0 (0%) | 1 (3%) |
| Weakness | 0 (0%) | 2 (13%) | 1 (8%) | 0 (0%) | 3 (8%) |
| Yawning | 1 (17%) | 1 (7%) | 1 (8%) | 0 (0%) | 3 (8%) |
| **Severity of adverse event during dosing** |  |  |  |  |  |
| Nil | 1 (17%) | 2 (13%) | 0 (0%) | 0 (0%) | 3 (8%) |
| Mild (an event tolerated by the participant, causing minimal discomfort and not interfering in everyday activities) | 5 (83%) | 13 (87%) | 12 (92%) | 5 (100%) | 35 (90%) |
| Moderate (an event sufficiently discomforting to interfere with normal everyday activities) | 0 (0%) | 0 (0%) | 1 (8%) | 0 (0%) | 1 (3%) |
| Serious adverse event during dosing |  |  |  |  |  |
| No | 6 (100%) | 15 (100%) | 13 (100%) | 5 (100%) | 39 (100%) |
| **Relatedness of adverse event to psilocybin during dosing** |  |  |  |  |  |
| not assessable | 0 (0%) | 1 (7%) | 0 (0%) | 0 (0%) | 1 (3%) |
| possibly related | 1 (17%) | 7 (47%) | 5 (38%) | 1 (20%) | 14 (36%) |
| probably related | 5 (83%) | 7 (47%) | 7 (54%) | 2 (40%) | 21 (54%) |
| definitely related | 0 (0%) | 0 (0%) | 1 (8%) | 2 (40%) | 3 (8%) |
| **Relatedness of adverse event to psilocybin during dosing** |  |  |  |  |  |
| No | 0 (0%) | 1 (7%) | 0 (0%) | 0 (0%) | 1 (3%) |
| Yes | 6 (100%) | 14 (93%) | 13 (100%) | 5 (100%) | 38 (97%) |
| **Adverse event or adverse reaction during dosing** |  |  |  |  |  |
| adverse event (an event which is unrelated to psilocybin) | 0 (0%) | 1 (7%) | 0 (0%) | 0 (0%) | 1 (3%) |
| adverse reaction (an event which is possibly/probably/definitely related to psilocybin) | 6 (100%) | 14 (93%) | 13 (100%) | 5 (100%) | 38 (97%) |
| **Significant Safety Issue during dosing** |  |  |  |  |  |
| No | 6 (100%) | 15 (100%) | 13 (100%) | 5 (100%) | 39 (100%) |
| **Adverse event post-dosing** | 4 (40%) | 4 (21%) | 11 (46%) | 6 (55%) | 25 (39%) |
| **Name of adverse event post-dosing** |  |  |  |  |  |
| Body ache | 0 (0%) | 1 (25%) | 0 (0%) | 1 (17%) | 2 (8%) |
| Drowsiness or sedation | 1 (25%) | 0 (0%) | 1 (9%) | 0 (0%) | 2 (8%) |
| Feeling cold inside | 0 (0%) | 0 (0%) | 1 (9%) | 0 (0%) | 1 (4%) |
| Feeling flushed in the face | 0 (0%) | 0 (0%) | 1 (9%) | 0 (0%) | 1 (4%) |
| Feeling run down | 1 (25%) | 0 (0%) | 0 (0%) | 0 (0%) | 1 (4%) |
| Headache | 1 (25%) | 2 (50%) | 3 (27%) | 2 (33%) | 8 (32%) |
| Hot and cold | 0 (0%) | 0 (0%) | 1 (9%) | 0 (0%) | 1 (4%) |
| Lightheadedness | 0 (0%) | 0 (0%) | 1 (9%) | 0 (0%) | 1 (4%) |
| Nausea with or without vomiting | 1 (25%) | 0 (0%) | 1 (9%) | 0 (0%) | 2 (8%) |
| Sound sensitivity | 0 (0%) | 0 (0%) | 0 (0%) | 1 (17%) | 1 (4%) |
| Tiredness | 0 (0%) | 1 (25%) | 1 (9%) | 1 (17%) | 3 (12%) |
| Warmth | 0 (0%) | 0 (0%) | 0 (0%) | 1 (17%) | 1 (4%) |
| Weakness | 0 (0%) | 0 (0%) | 1 (9%) | 0 (0%) | 1 (4%) |
| **Severity of adverse event post-dosing** |  |  |  |  |  |
| Nil | 2 (50%) | 0 (0%) | 0 (0%) | 1 (17%) | 3 (12%) |
| Mild (an event tolerated by the participant, causing minimal discomfort and not interfering in everyday activities) | 2 (50%) | 4 (100%) | 6 (55%) | 5 (83%) | 17 (68%) |
| Moderate (an event sufficiently discomforting to interfere with normal everyday activities) | 0 (0%) | 0 (0%) | 4 (36%) | 0 (0%) | 4 (16%) |
| Severe (an event that prevents normal everyday activities) | 0 (0%) | 0 (0%) | 1 (9%) | 0 (0%) | 1 (4%) |
| **Serious adverse event post-dosing** |  |  |  |  |  |
| No | 4 (100%) | 4 (100%) | 11 (100%) | 6 (100%) | 25 (100%) |
| **Relatedness of adverse event to psilocybin post-dosing** |  |  |  |  |  |
| not assessable | 1 (25%) | 0 (0%) | 0 (0%) | 0 (0%) | 1 (4%) |
| possibly related | 2 (50%) | 4 (100%) | 10 (91%) | 6 (100%) | 22 (88%) |
| probably related | 1 (25%) | 0 (0%) | 1 (9%) | 0 (0%) | 2 (8%) |
| **Relatedness of adverse event to psilocybin post-dosing** |  |  |  |  |  |
| No | 1 (25%) | 0 (0%) | 0 (0%) | 0 (0%) | 1 (4%) |
| Yes | 3 (75%) | 4 (100%) | 11 (100%) | 6 (100%) | 24 (96%) |
| **Adverse event or adverse reaction post-dosing** |  |  |  |  |  |
| adverse event (an event which is unrelated to psilocybin) | 1 (25%) | 0 (0%) | 0 (0%) | 0 (0%) | 1 (4%) |
| adverse reaction (an event which is possibly/probably/definitely related to psilocybin) | 3 (75%) | 4 (100%) | 11 (100%) | 6 (100%) | 24 (96%) |
| **Significant Safety Issue post-dosing** |  |  |  |  |  |
| No | 4 (100%) | 4 (100%) | 11 (100%) | 6 (100%) | 25 (100%) |

Values refer to the number (%) of adverse events.

### Vital Signs

**Table S2.2: Vital Signs**

|  | **5mg** | **10mg** | **15mg** | **20mg** |
| --- | --- | --- | --- | --- |
| **Systolic blood pressure (mmHg)** | N=7 | N=12 | N=11 | N=6 |
| Baseline (pre-dosing) | 125.0 (116.0-132.0) | 124.0 (119.0-135.5) | 119.0 (113.0-138.0) | 118.0 (112.0-132.0) |
| 0.5 hour post-dosing | 123.5 (114.0-131.0) | 130.0 (122.0-140.5) | 130.0 (123.0-141.0) | 127.0 (117.0-160.0) |
| 0.5 hour post-dosing (change from baseline) | -6.5 (-11.0-3.0) | 4.0 (0.0-11.5) | 8.0 (-7.0-15.0) | 6.5 (-6.0-15.0) |
| 1 hour post-dosing | 133.0 (125.0-133.0) | 134.0 (125.5-142.5) | 132.0 (123.0-147.0) | 133.0 (133.0-150.0) |
| 1 hour post-dosing (change from baseline) | 3.0 (1.0-8.0) | 4.5 (1.5-13.0) | 4.0 (-1.0-15.0) | 5.0 (2.0-14.0) |
| 3 hours post-dosing | 124.0 (120.0-134.0) | 139.0 (122.0-148.0) | 135.0 (133.0-145.0) | 133.0 (123.0-140.0) |
| 3 hours post-dosing (change from baseline) | 2.0 (-5.0-5.0) | 7.0 (3.0-17.0) | 10.0 (5.0-14.0) | 8.0 (-5.0-14.0) |
| 5 hours post-dosing | 128.0 (120.0-134.0) | 125.5 (118.5-139.5) | 131.0 (121.0-141.0) | 126.0 (125.0-146.0) |
| 5 hours post-dosing (change from baseline) | 0.0 (-7.0-7.0) | 0.0 (-6.0-11.5) | 2.0 (-3.0-6.0) | 3.5 (-4.0-7.0) |
| **Diastolic blood pressure (mmHg)** | 7 (100%) | 12 (100%) | 11 (100%) | 6 (100%) |
| Baseline (pre-dosing) | 82.0 (79.0-86.0) | 80.0 (77.5-83.5) | 82.0 (78.0-84.0) | 82.0 (78.0-89.0) |
| 0.5 hour post-dosing | 81.5 (78.0-85.0) | 86.5 (82.0-93.5) | 85.0 (82.0-94.0) | 88.0 (75.0-90.0) |
| 0.5 hour post-dosing (change from baseline) | 0.5 (-4.0-3.0) | 4.5 (-0.5-11.0) | 3.0 (0.0-15.0) | 0.5 (-2.0-15.0) |
| 1 hour post-dosing | 88.0 (83.0-92.0) | 88.0 (83.5-90.5) | 94.0 (84.0-99.0) | 89.0 (85.0-94.0) |
| 1 hour post-dosing (change from baseline) | 3.5 (0.0-7.0) | 6.5 (2.5-11.0) | 10.0 (6.0-18.0) | 14.0 (-4.0-18.0) |
| 3 hours post-dosing | 78.0 (77.0-93.0) | 86.0 (78.0-94.0) | 90.0 (81.0-95.0) | 86.0 (79.0-97.0) |
| 3 hours post-dosing (change from baseline) | 4.0 (-7.0-5.0) | 6.0 (-4.0-11.0) | 4.0 (0.0-16.0) | 4.0 (-6.0-14.0) |
| 5 hours post-dosing | 87.0 (77.0-92.0) | 82.0 (74.5-87.5) | 81.0 (73.0-94.0) | 81.0 (72.0-91.0) |
| 5 hours post-dosing (change from baseline) | 5.0 (-5.0-7.0) | 2.0 (-8.5-7.0) | 1.0 (-5.0-6.0) | 1.5 (-12.0-7.0) |
| **Pulse rate (per minute)** | 7 (100%) | 12 (100%) | 11 (100%) | 6 (100%) |
| Baseline (pre-dosing) | 75.0 (55.0-88.0) | 79.5 (68.5-83.0) | 80.0 (64.0-83.0) | 75.0 (65.0-80.0) |
| 0.5 hour post-dosing | 63.0 (49.0-65.0) | 78.0 (68.5-89.0) | 76.0 (65.0-82.0) | 78.0 (64.0-79.0) |
| 0.5 hour post-dosing (change from baseline) | -11.5 (-13.0-0.0) | -3.0 (-8.5-12.5) | 0.0 (-4.0-11.0) | -4.5 (-6.0--3.0) |
| 1 hour post-dosing | 65.5 (50.0-74.0) | 79.5 (67.5-86.0) | 75.0 (70.0-80.0) | 76.0 (76.0-80.0) |
| 1 hour post-dosing (change from baseline) | -6.0 (-17.0--1.0) | 4.0 (-8.0-8.0) | 2.0 (-6.0-6.0) | -6.0 (-10.0--2.0) |
| 3 hours post-dosing | 62.0 (56.0-78.0) | 79.0 (67.0-96.0) | 76.0 (70.0-80.0) | 80.0 (74.0-82.0) |
| 3 hours post-dosing (change from baseline) | -4.0 (-10.0-2.0) | 7.0 (-7.0-16.0) | 5.0 (-9.0-10.0) | 2.0 (-8.0-6.0) |
| 5 hours post-dosing | 70.0 (64.0-84.0) | 87.0 (71.0-90.0) | 80.0 (74.0-88.0) | 89.5 (85.0-92.0) |
| 5 hours post-dosing (change from baseline) | -2.0 (-5.0-5.0) | 5.5 (-1.5-11.5) | -1.0 (-4.0-8.0) | 6.5 (3.0-9.0) |
| **Oxygen saturation (% at room air)** | 7 (100%) | 12 (100%) | 11 (100%) | 6 (100%) |
| Baseline (pre-dosing) | 99.0 (97.0-99.0) | 98.0 (98.0-98.5) | 98.0 (98.0-99.0) | 96.0 (95.0-98.0) |
| 0.5 hour post-dosing | 99.0 (98.0-99.0) | 98.5 (97.0-99.0) | 98.0 (98.0-99.0) | 98.0 (97.0-98.0) |
| 0.5 hour post-dosing (change from baseline) | 1.0 (0.0-1.0) | 0.0 (-1.0-1.5) | 0.0 (0.0-1.0) | 0.0 (-1.0-0.0) |
| 1 hour post-dosing | 98.5 (98.0-99.0) | 98.0 (97.0-99.0) | 98.0 (98.0-99.0) | 98.0 (97.0-98.0) |
| 1 hour post-dosing (change from baseline) | 1.0 (-1.0-1.0) | 0.5 (-1.5-1.0) | 0.0 (-1.0-1.0) | 0.0 (-1.0-0.0) |
| 3 hours post-dosing | 98.0 (97.0-99.0) | 98.0 (95.0-98.0) | 98.0 (97.0-98.0) | 97.0 (97.0-98.0) |
| 3 hours post-dosing (change from baseline) | 0.0 (-1.0-1.0) | -1.0 (-3.0-0.0) | 0.0 (-1.0-1.0) | 0.0 (-1.0-0.0) |
| 5 hours post-dosing | 98.0 (98.0-99.0) | 98.0 (95.5-98.0) | 97.0 (96.0-98.0) | 96.5 (96.0-98.0) |
| 5 hours post-dosing (change from baseline) | 0.0 (-1.0-0.0) | -1.0 (-2.5-0.5) | -1.0 (-2.0-0.0) | -1.0 (-2.0--1.0) |

Data are presented as median (interquartile range) for continuous measures, and n (%) for categorical measures.

### Administrative Information

**Author Names:** Dr Chiranth Bhagavan, Professor Olivia Carter, Dr Glenn Nielsen, Professor David Berlowitz, Ms Sara Issak, Associate Professor Sabine Braat, Dr Sophie Zaloumis, Mr Zachary Attard, Ms Gina Oliver, Ms Deanne Mayne, Dr James Rucker, Dr Matthew Butler, Dr Orwa Dandash, Dr Alexander Bryson, and Professor Richard A. Kanaan.

**Corresponding Author:** Dr Chiranth Bhagavan, Department of Psychiatry, University of Melbourne, Austin Health, 145 Studley Rd, Heidelberg VIC 3084, Australia;; phone: +61 493 766 597.
