## Supplementary Information 2 for "A Randomised, Triple-Blind, Dose-Finding Study of the Impact of Psilocybin on Motor Function in Healthy Participants"

### Supplementary Information 3: Individual Profiles

### Motor function

#### Combined de Morton Mobility Index and Functional Movement Exploration (% Maximum)

“demmi_fnd_extension_total_perc”

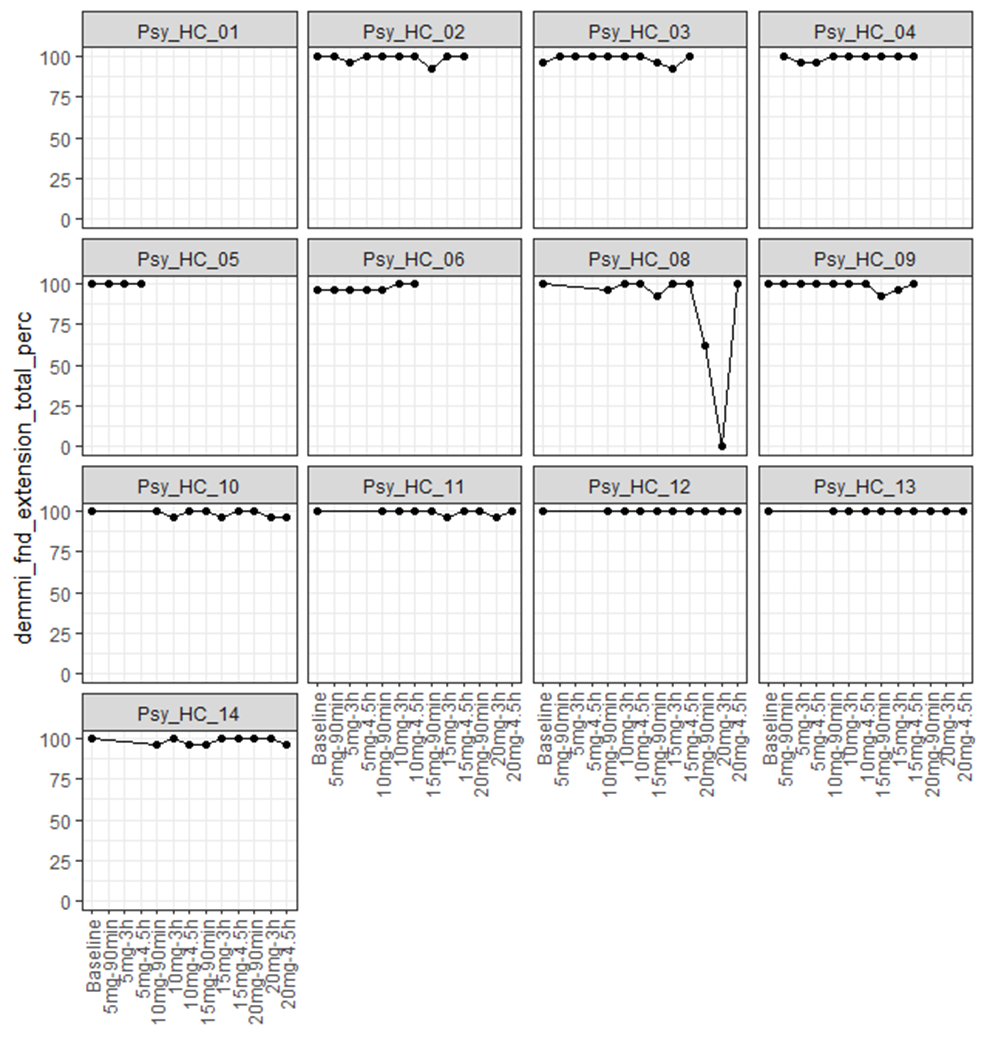

**Figure S3.1:** Individual profiles for demmi_fnd_extension_total_perc.

#### Action Research Arm Test

“arat_total”

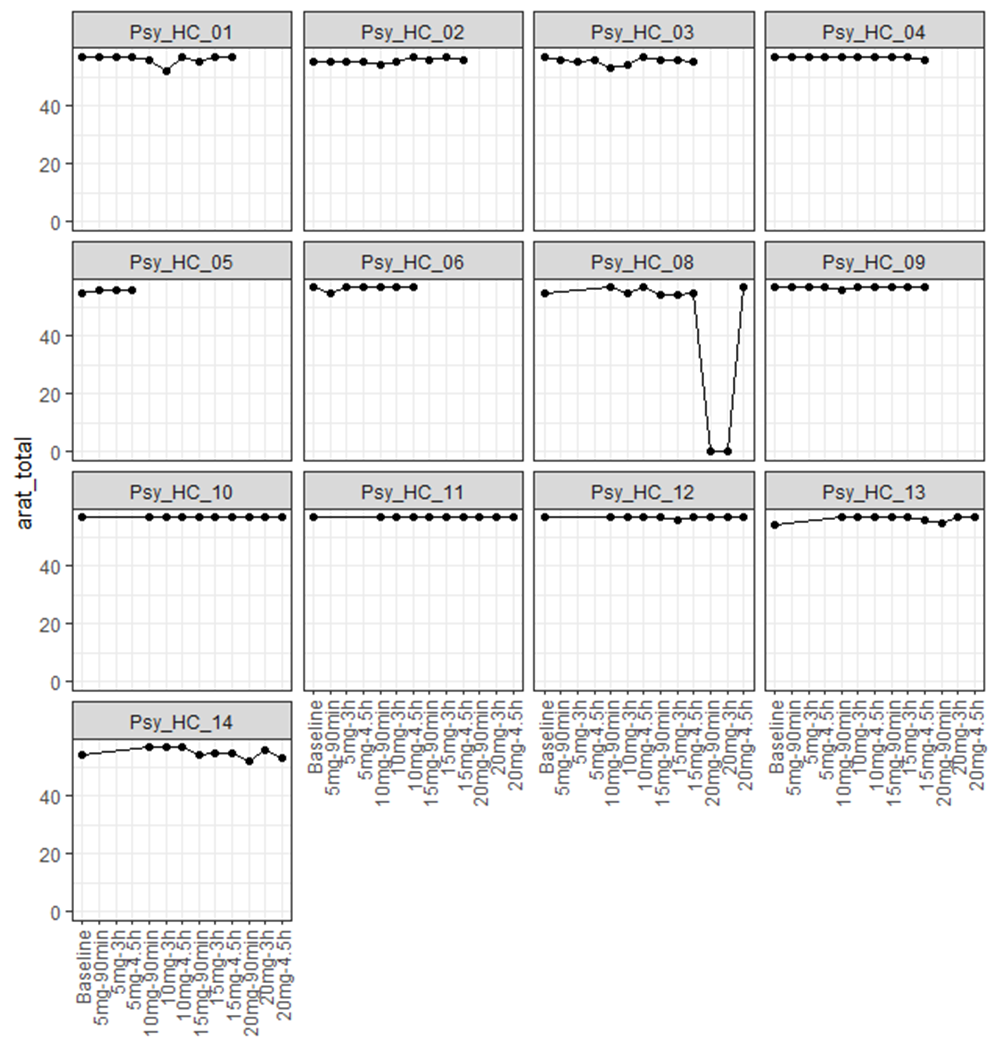

**Figure S3.2:** Individual profiles for arat_total.

#### Box and Block (Original)

“boxblock_std_score”

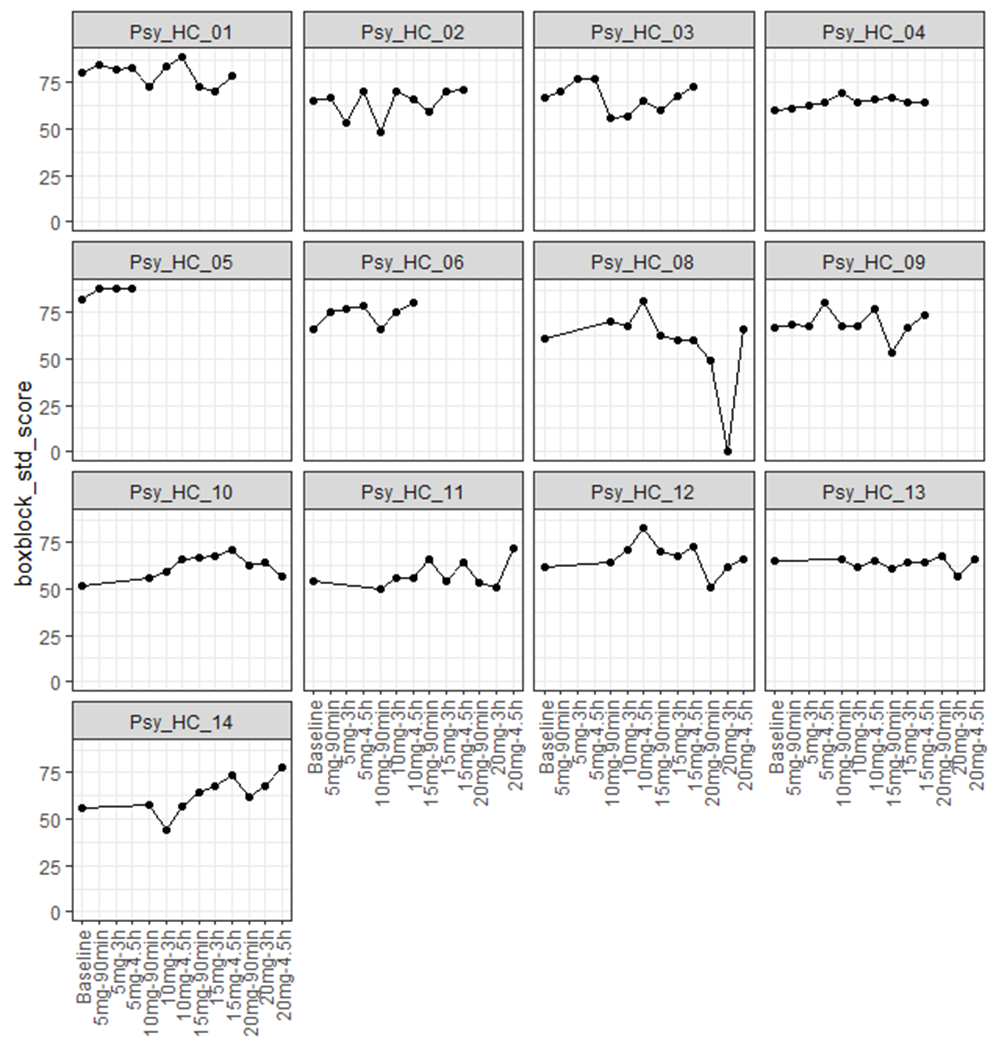

**FigureS3.3:** Individual profiles for boxblock_std_score.

#### Box and Block (Modified)

“boxblock_mod_score”

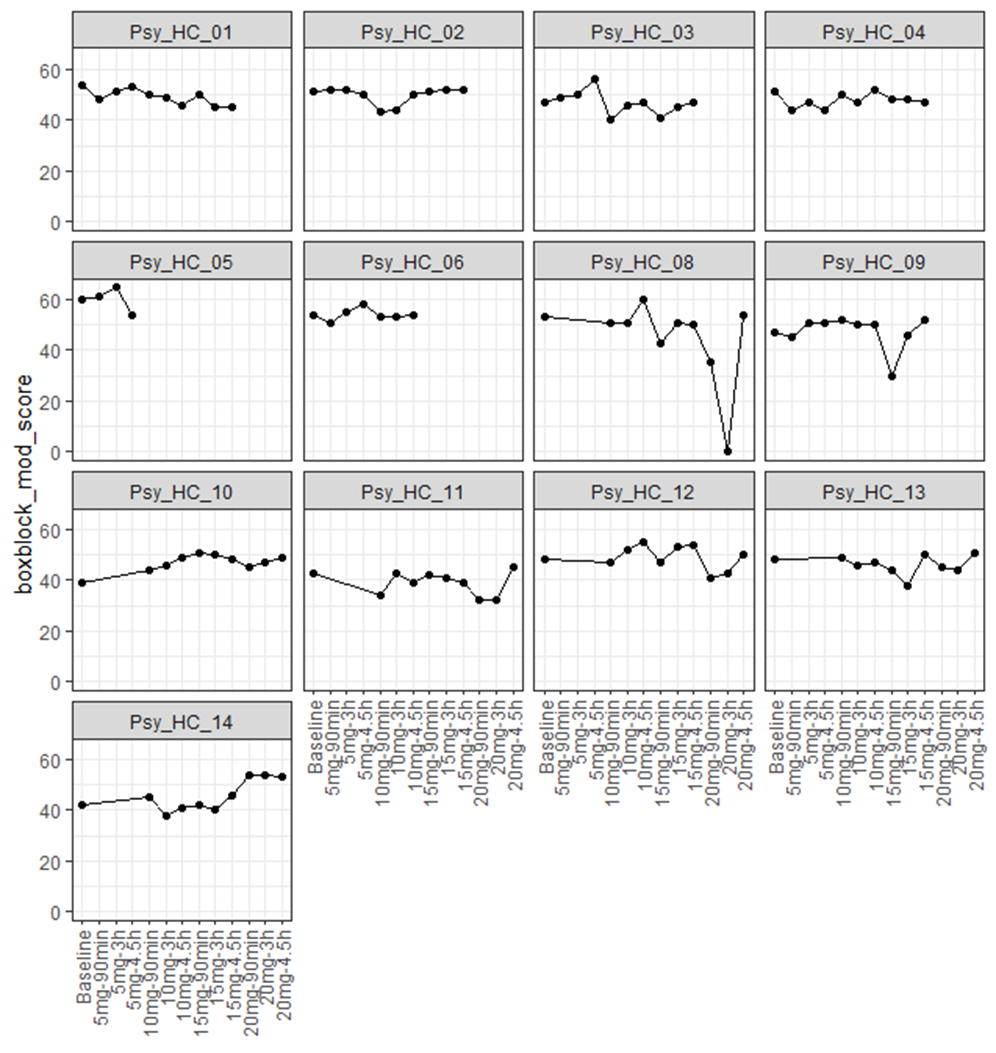

**Figure S3.4:** Individual profiles for boxblock_mod_score.

#### Digit Symbol Substitution Test

“dsst_score”

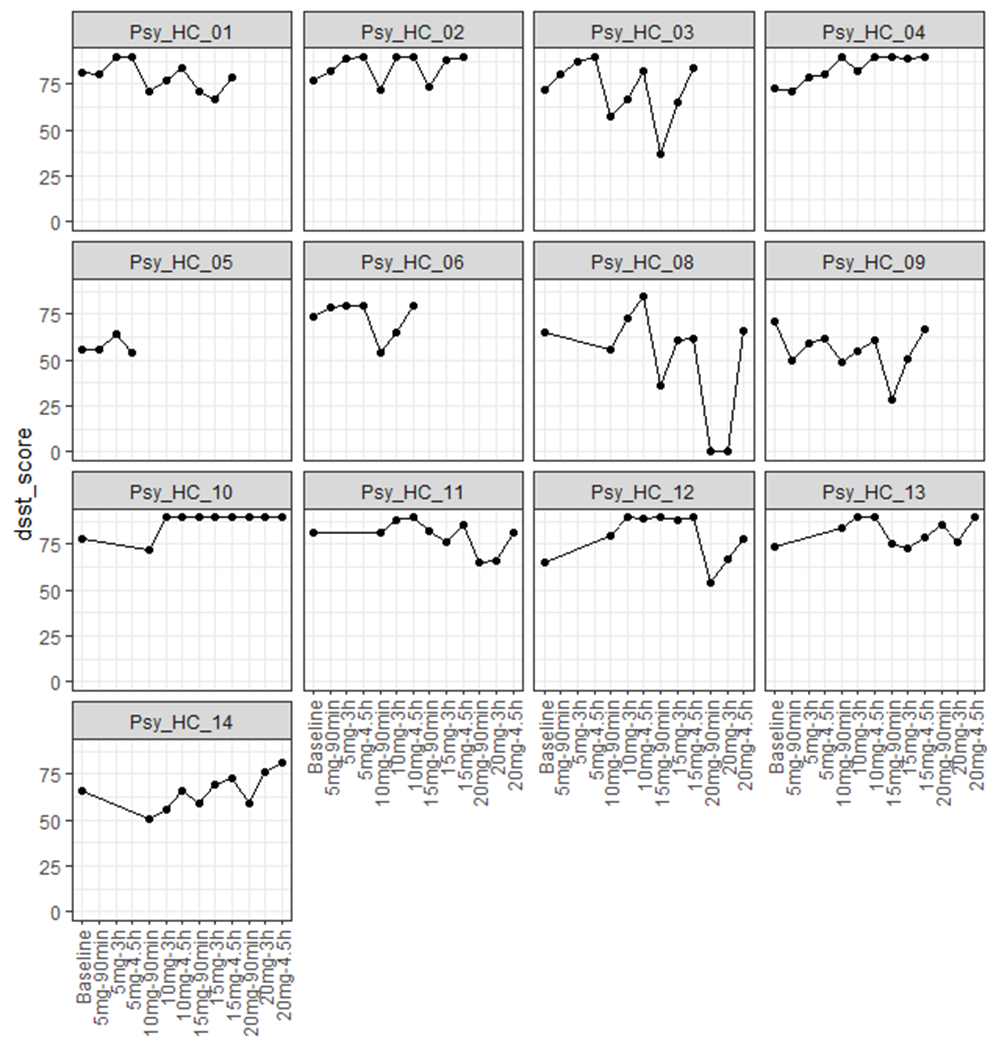

**Figure S3.5:** Individual profiles for dsst_score.

#### Reaction Time Ruler Drop Test (Mean)

“rt_ruler_mean”

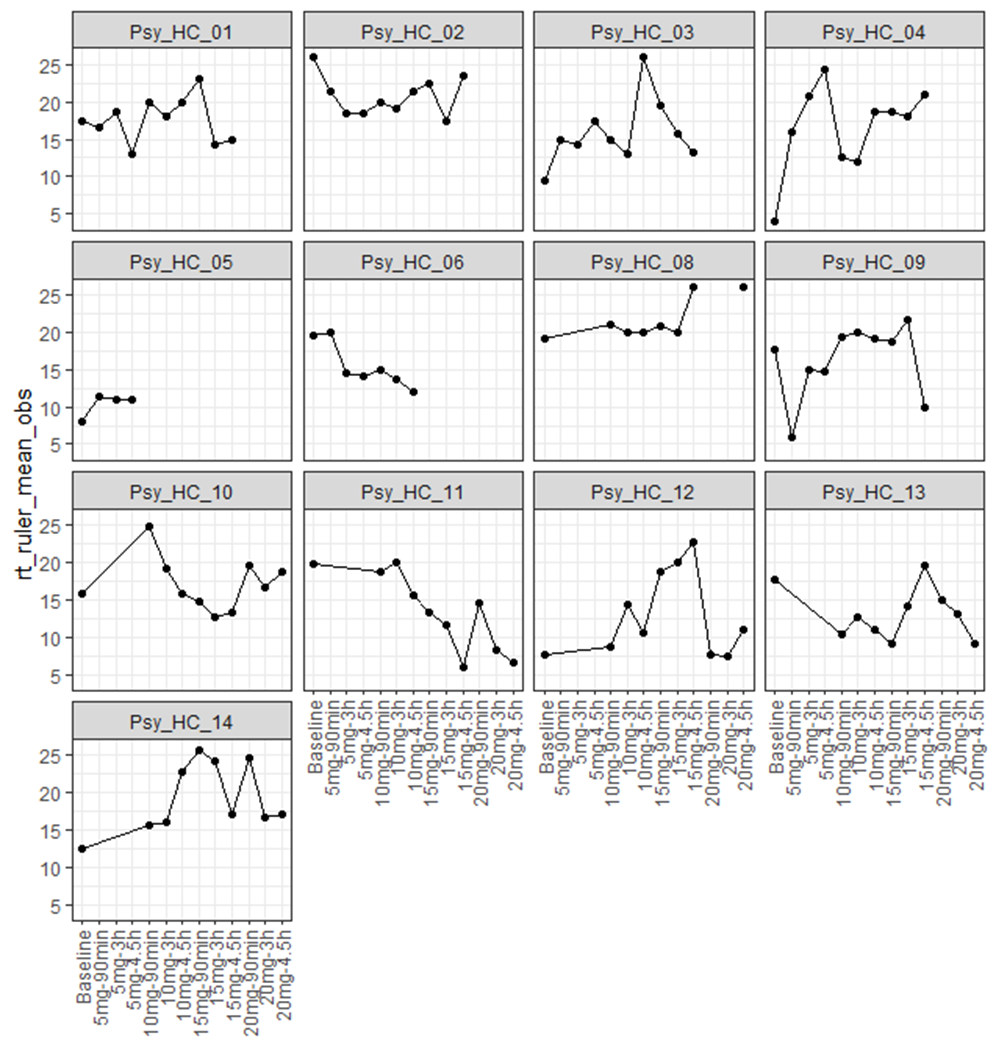

**Figure S3.6:** Individual profiles for rt_ruler_mean.

### Ego-Dissolution Inventory

#### Ego-Dissolution Inventory (Mean)

“edi_mean”

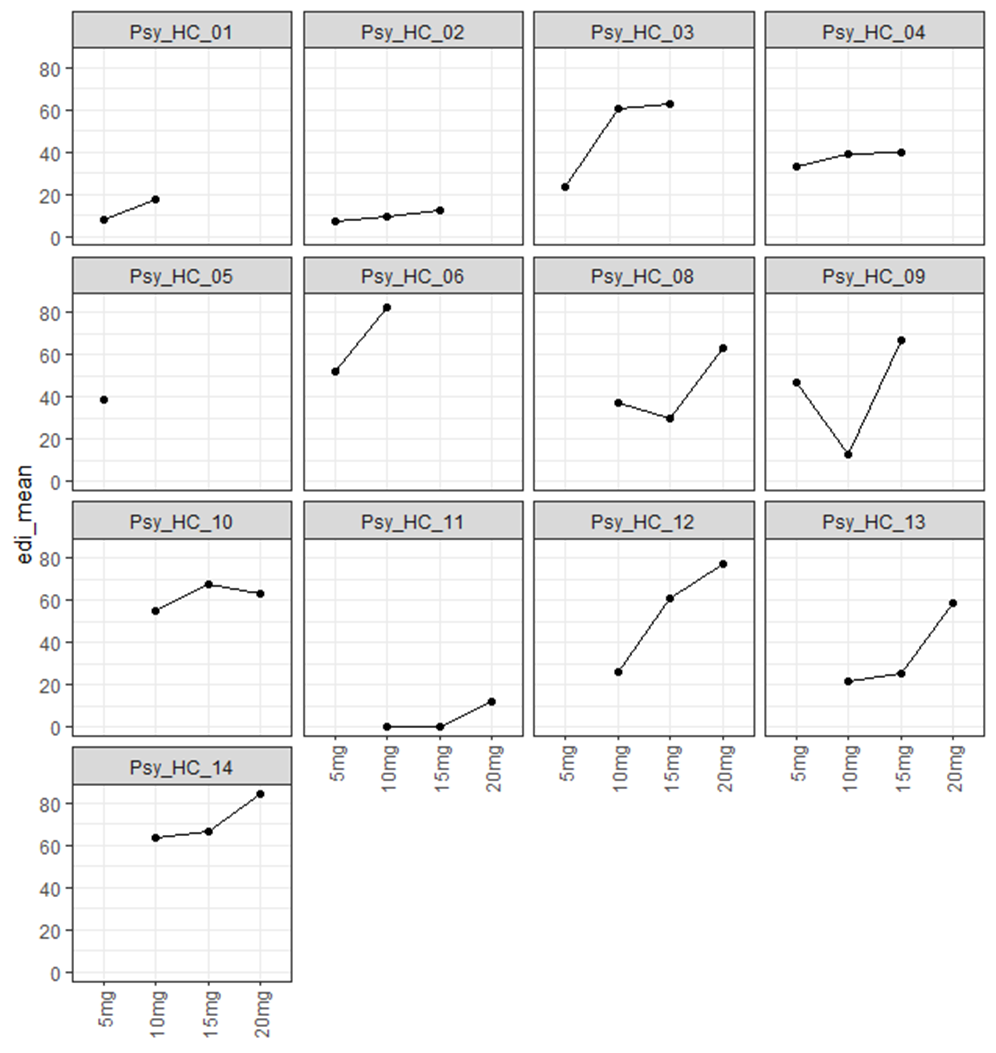

**Figure S3.7:** Individual profiles for edi_mean.

#### Q1: Dissolution of self

“edi_1”

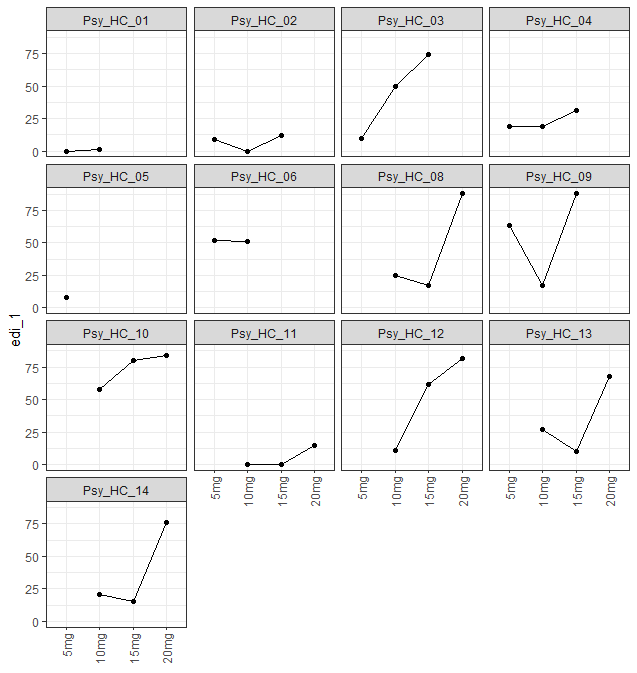

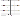
***Figure S3.8:*** *Individual profiles for edi_1.*

#### Q2: Feeling one with the universe

“edi_2”

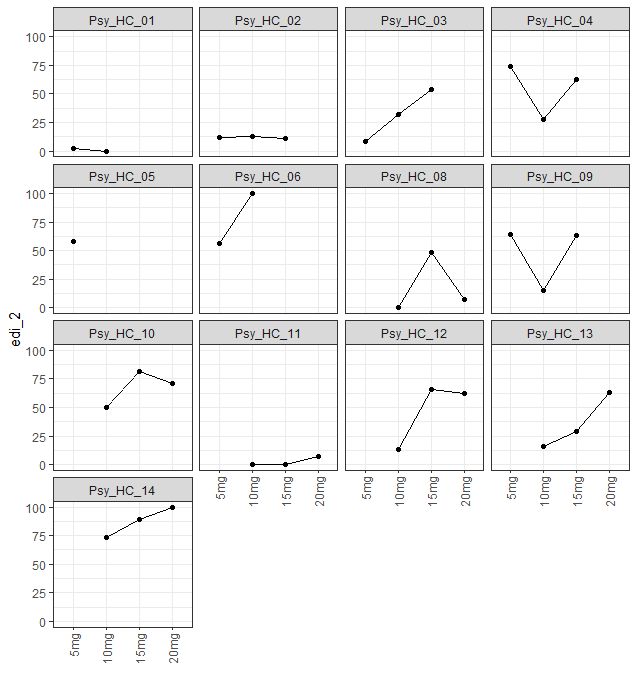

**Figure** **S3.9:** Individual profiles for edi_2.

#### Q3: Sense of union with others

“edi_3”

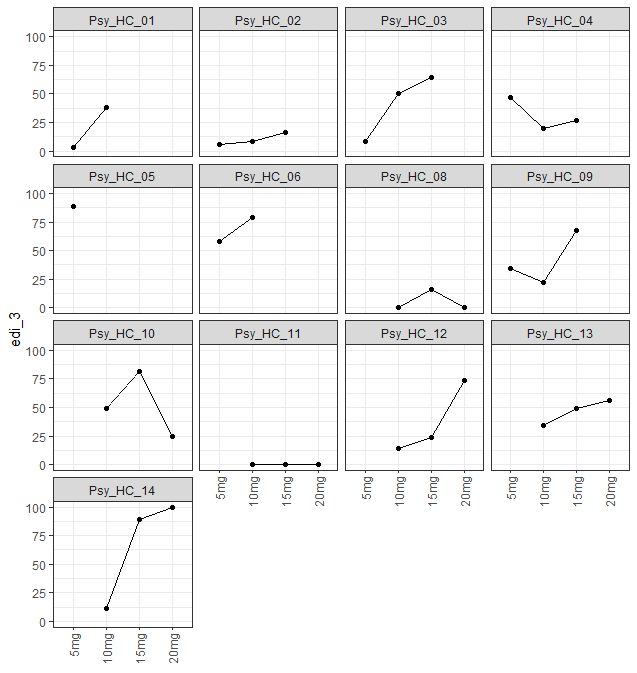

**Figure S3.10:** Individual profiles for edi_3.

#### Q4: Decrease in sense of self-importance

“edi_4”

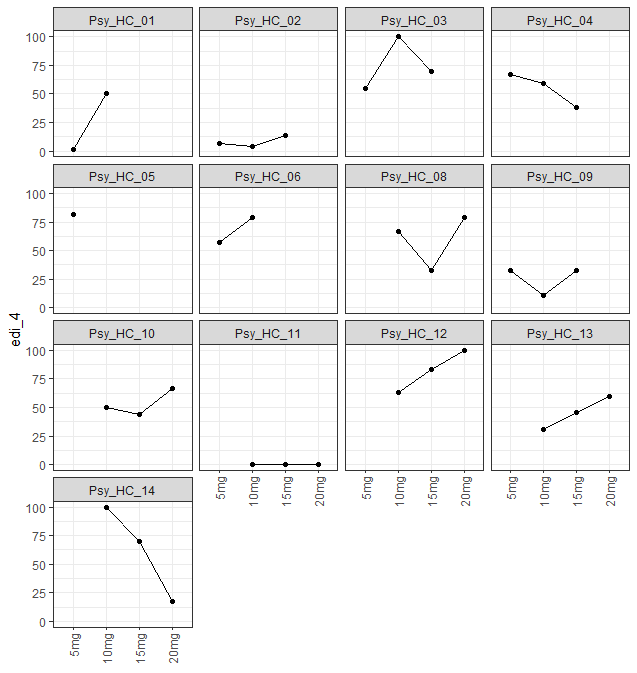

**Figure S3.11:** Individual profiles for edi_4.

#### Q5: Disintegration of self or ego

“edi_5”

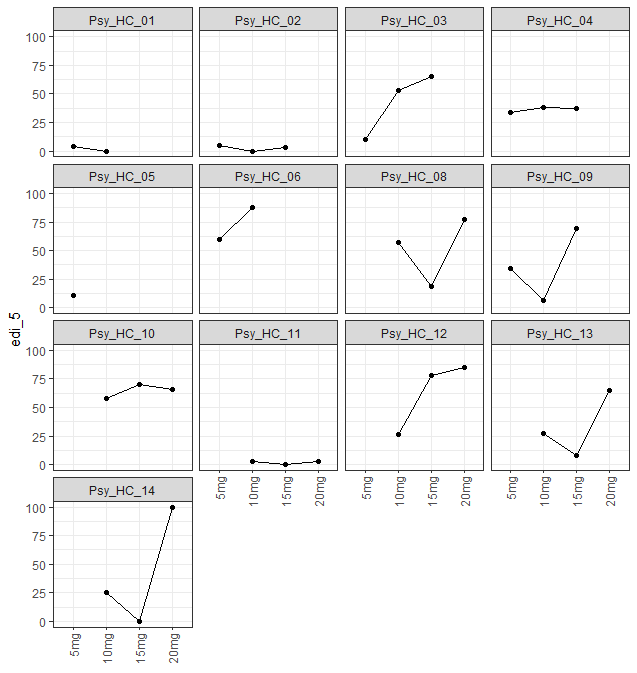

**Figure S3.12:** Individual profiles for edi_5.

#### Q6: Less absorbed with own issues/concerns

“edi_6”

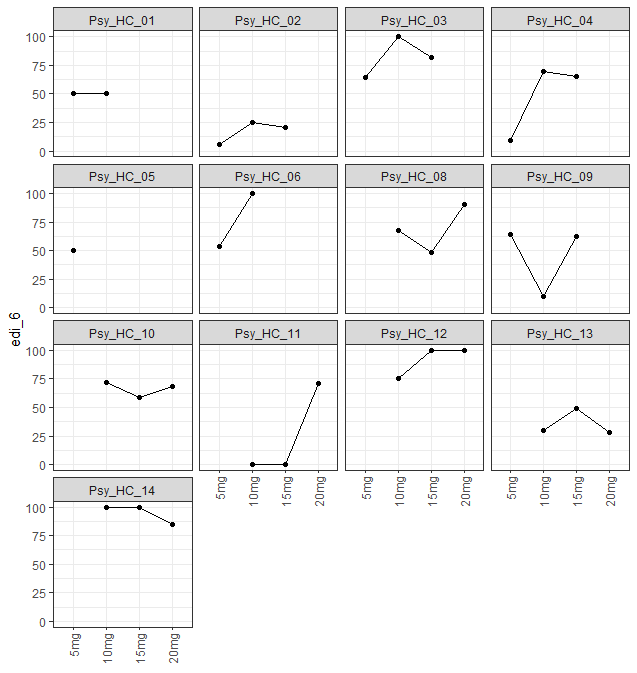

**Figure S3.13:** Individual profiles for edi_6.

#### Q7: Loss of sense of ego

“edi_7”

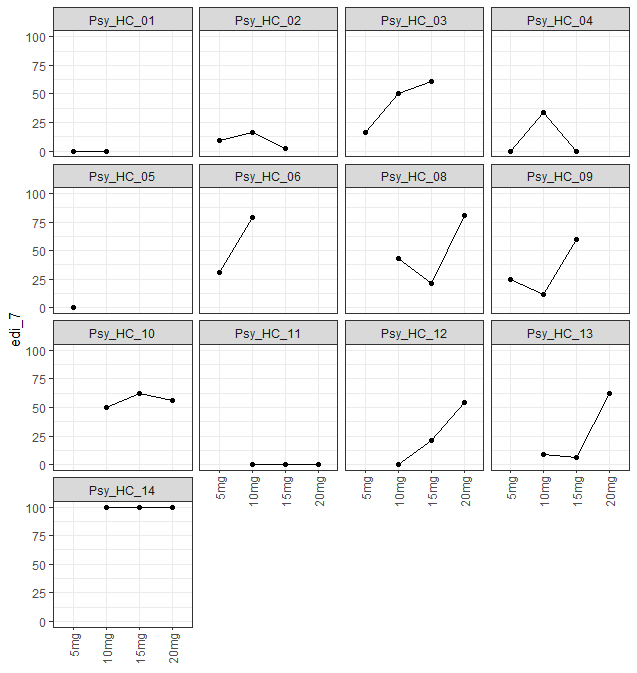

**Figure S3.14:** Individual profiles for edi_7.

#### Q8: All notion of self and identity dissolved away

“edi_8”

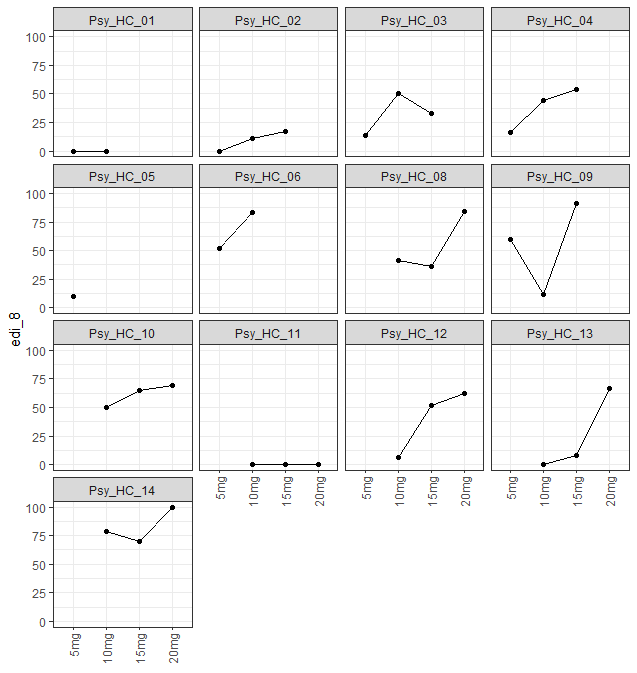

**Figure S3.15:** Individual profiles for edi_8.

### 5-Dimensional Altered States of Consciousness

#### 5-Dimensional Altered States of Consciousness (Mean)

“d5_asc_mean”

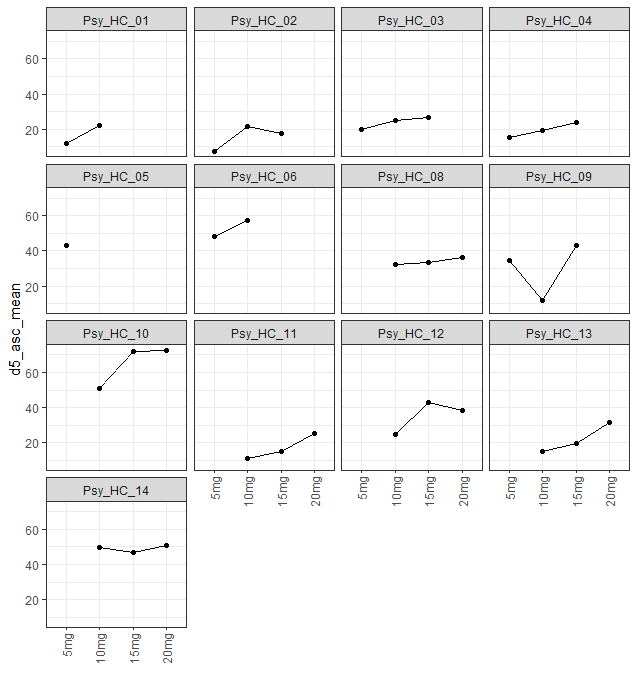

**Figure S3.16:** Individual profiles for d5_asc_mean.

#### Experience of Unity

“Experience_of_Unity”

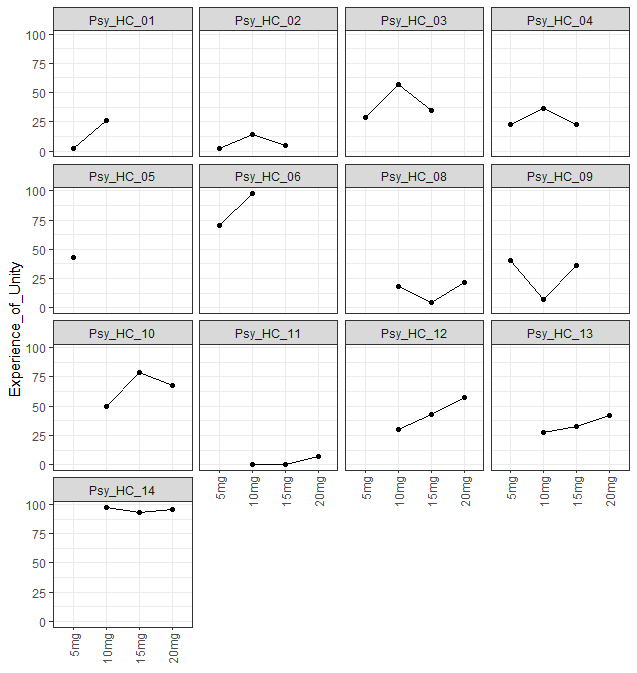

**Figure S3.17:** Individual profiles for Experience_of_Unity.

#### Spiritual Experience

“Spiritual_Experience”

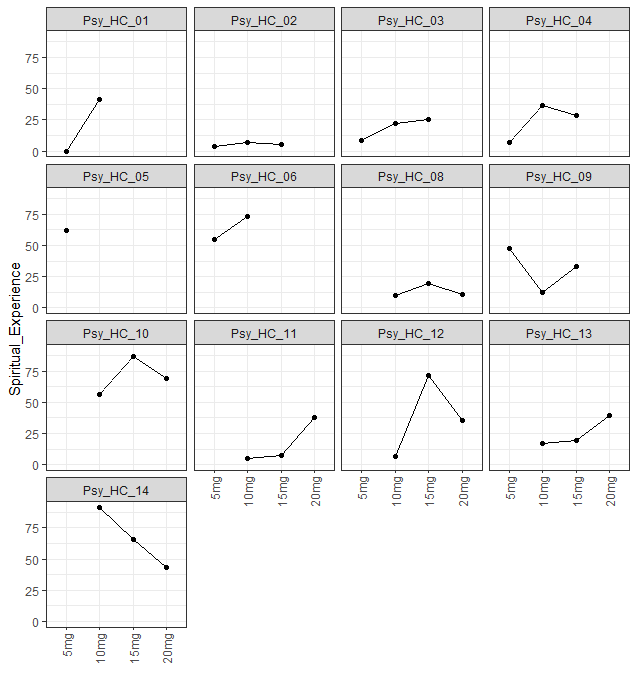

**Figure S3.18:** Individual profiles for Spiritual_Experience.

#### Blissful State

“Blissful_State”

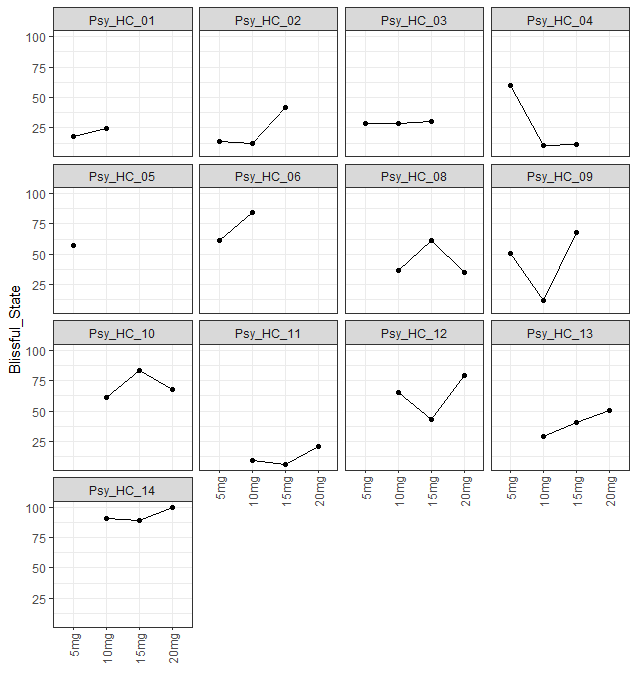

**Figure S3.19:** Individual profiles for Blissful_State.

#### Insightfulness

“Insightfulness”

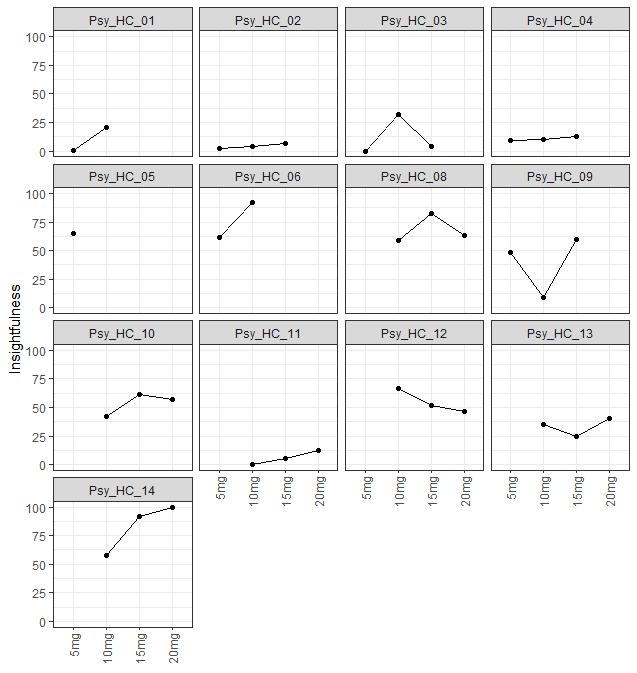

**Figure S3.20:** Individual profiles for Insightfulness.

#### Disembodiment

“Disembodiment”

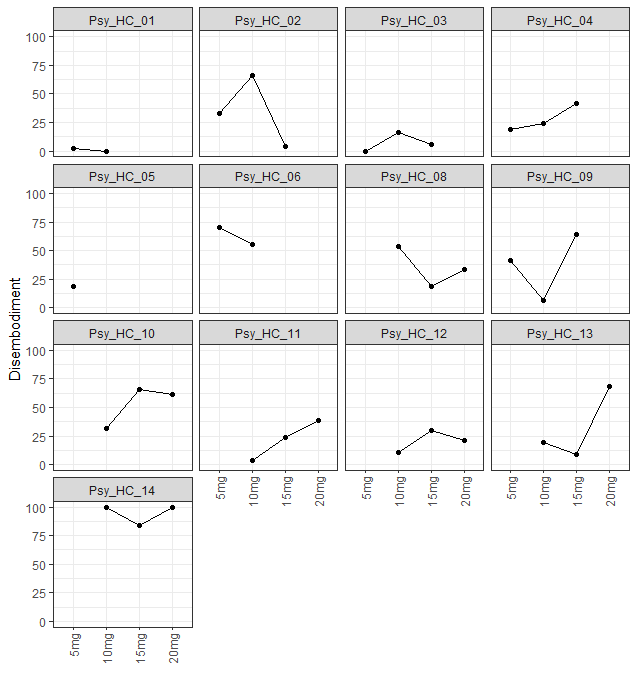

**Figure S3.21:** Individual profiles for Disembodiment.

#### Impaired Control and Cognition

“Impaired_Control_and_Cognition”

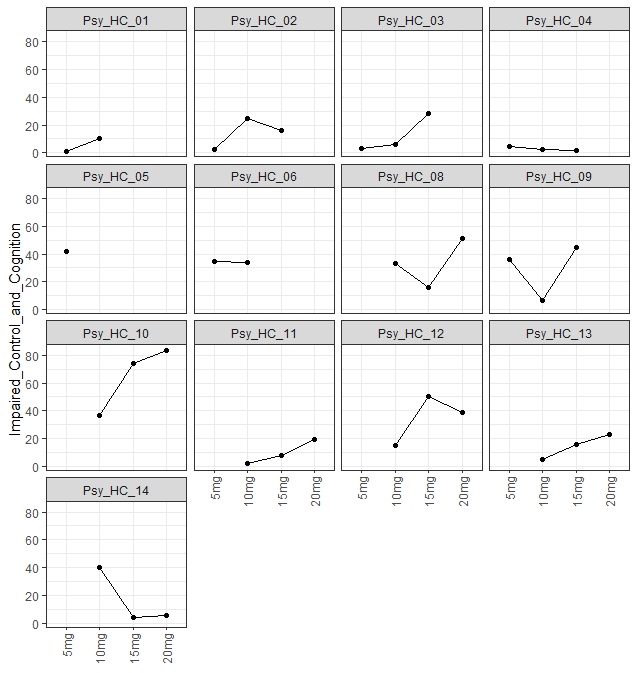

**Figure S3.22:** Individual profiles for Impaired_Control_and_Cognition.

#### Anxiety

“Anxiety”

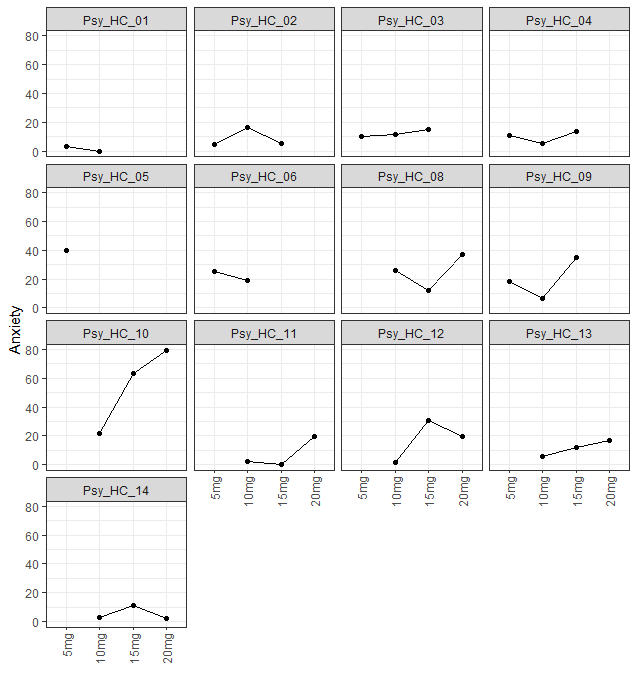

**Figure S3.23:** Individual profiles for Anxiety.

#### Complex Imagery

“Complex_Imagery”

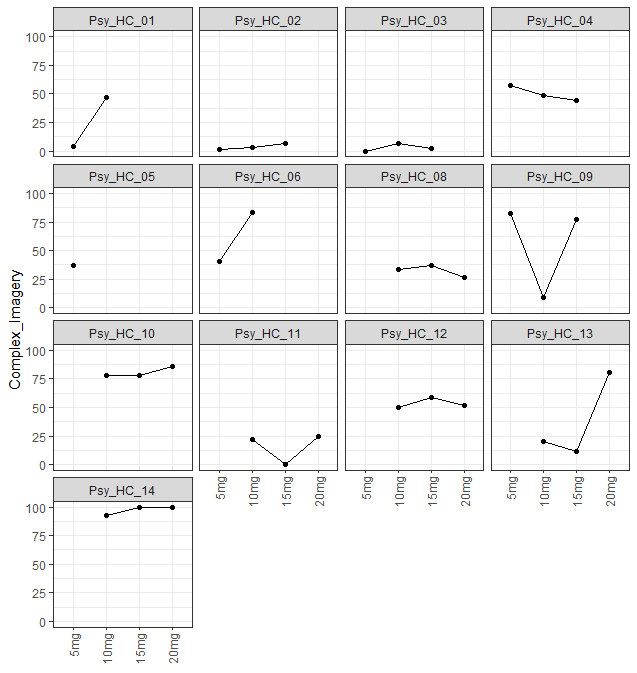

**Figure S3.24:** Individual profiles for Complex_Imagery.

#### Elementary Imagery

“Elementary_Imagery”

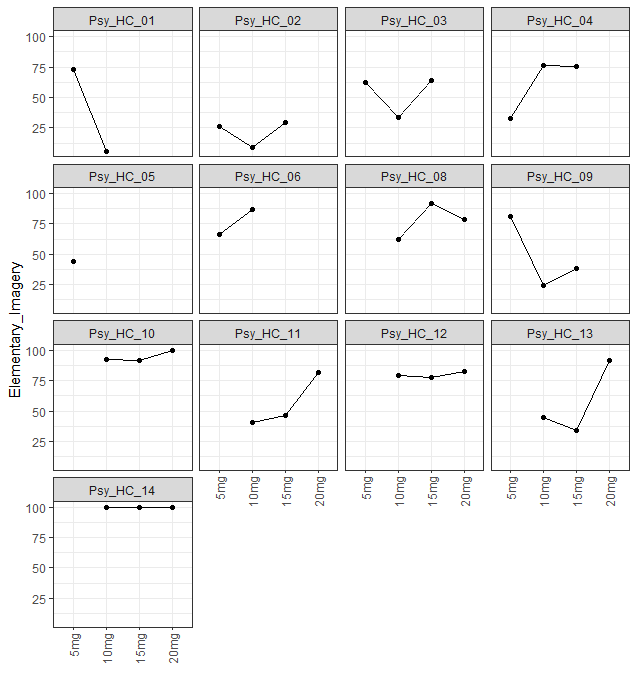

**Figure S3.25:** Individual profiles for Elementary_Imagery.

#### Audio Visual Synesthesiae

“Audio_Visual_Synesthesiae”

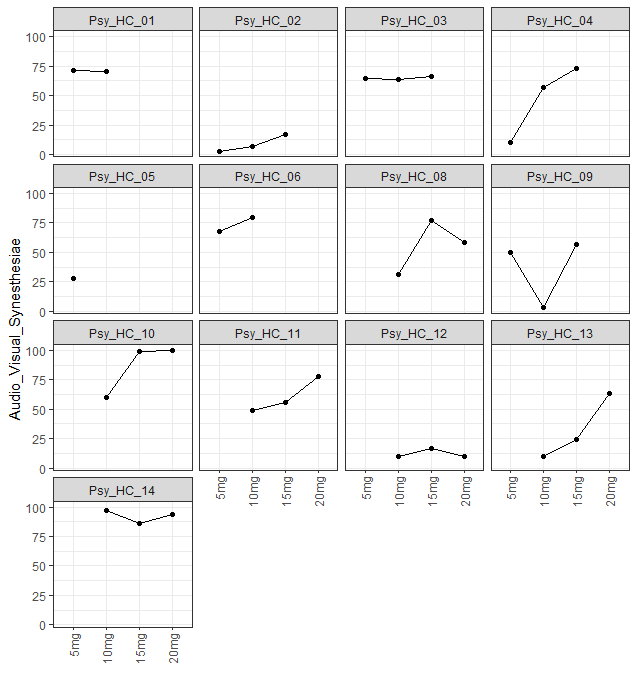

**Figure S3.26:** Individual profiles for Audio_Visual_Synesthesiae.

#### Changed Meaning of Percepts

“Changed_Meaning_of_Percepts”

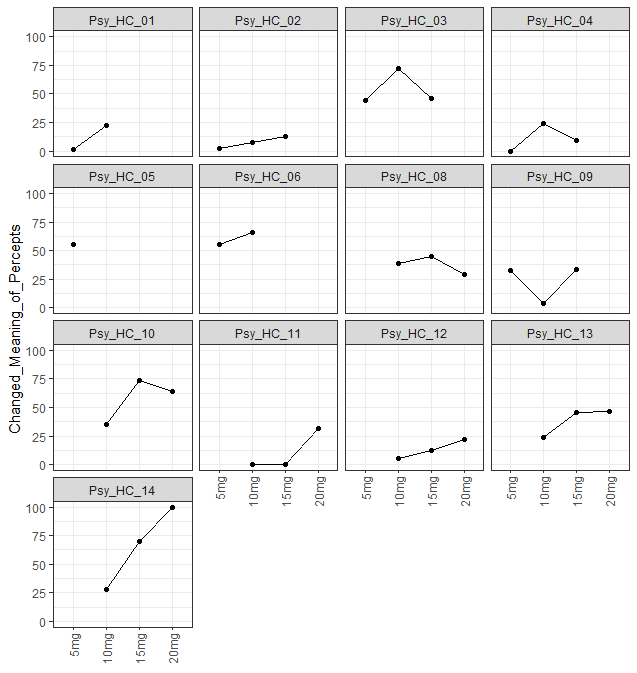

**Figure S3.27:** Individual profiles for Changed_Meaning_of_Percepts.

### Administrative Information

**Author Names:** Dr Chiranth Bhagavan, Professor Olivia Carter, Dr Glenn Nielsen, Professor David Berlowitz, Ms Sara Issak, Associate Professor Sabine Braat, Dr Sophie Zaloumis, Mr Zachary Attard, Ms Gina Oliver, Ms Deanne Mayne, Dr James Rucker, Dr Matthew Butler, Dr Orwa Dandash, Dr Alexander Bryson, and Professor Richard A. Kanaan.

**Corresponding Author:** Dr Chiranth Bhagavan, Department of Psychiatry, University of Melbourne, Austin Health, 145 Studley Rd, Heidelberg VIC 3084, Australia;; phone: +61 493 766 597.
