## Supplementary Information 4 for "A Randomised, Triple-Blind, Dose-Finding Study of the Impact of Psilocybin on Motor Function in Healthy Participants"

### Supplementary Information 5: Post-Hoc Analysis

### Motor function

#### Combined de Morton Mobility Index and Functional Movement Exploration (% Maximum)

“demmi_fnd_extension_total_perc”

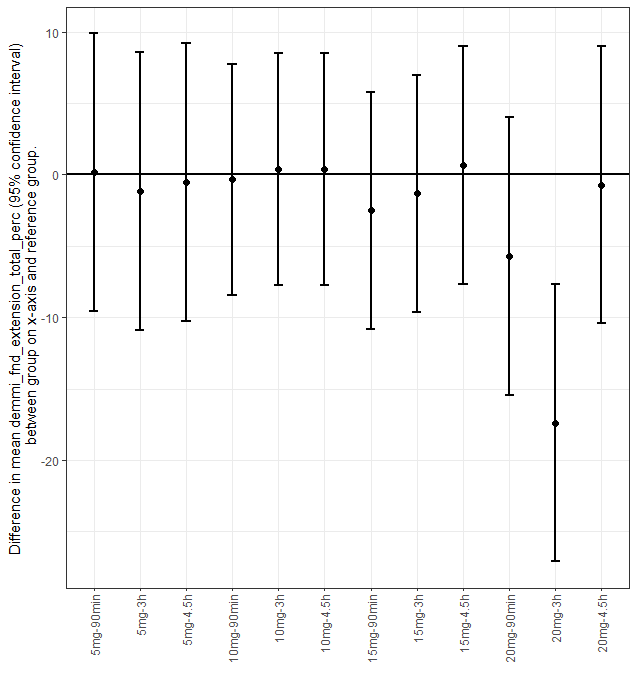

**Figure S5.1:** Caterpillar plot for demmi_fnd_extension_total_perc.

#### Action Research Arm Test

“arat_total”

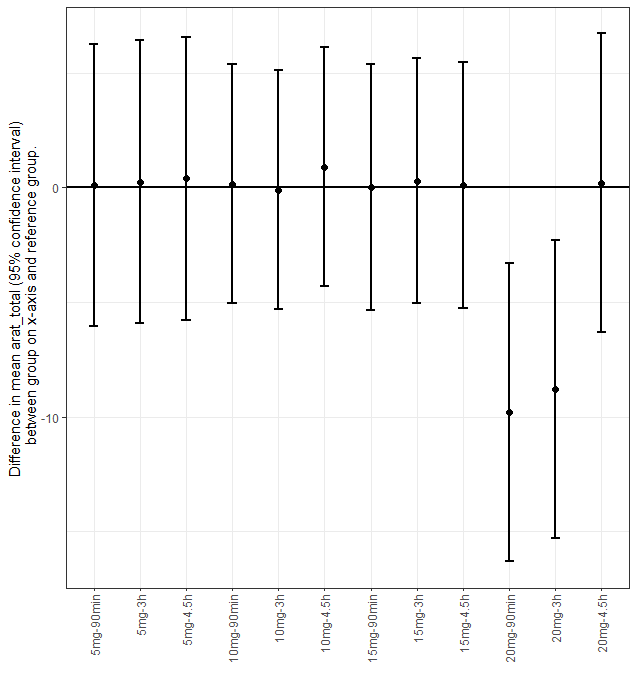

**Figure S5.2:** Caterpillar plot for arat_total.

#### Box and Block (Original)

“boxblock_std_score”

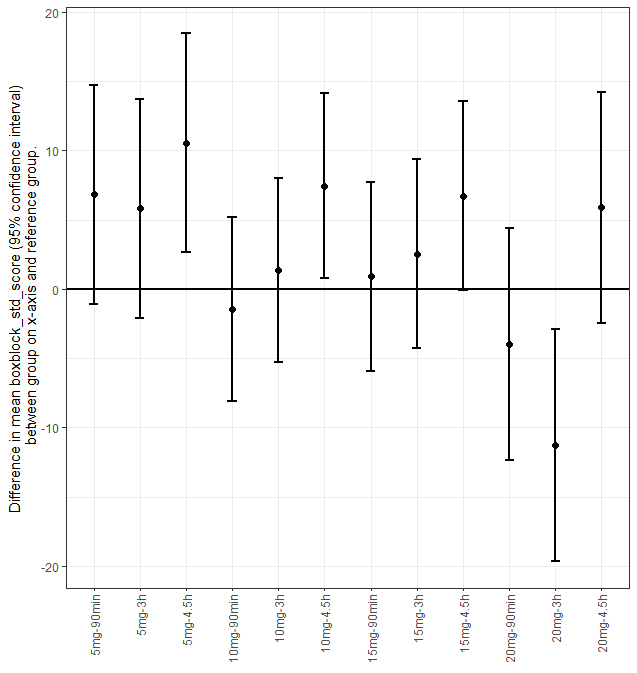

**FigureS5.3:** Caterpillar plot for boxblock_std_score.

#### Box and Block (Modified)

“boxblock_mod_score”

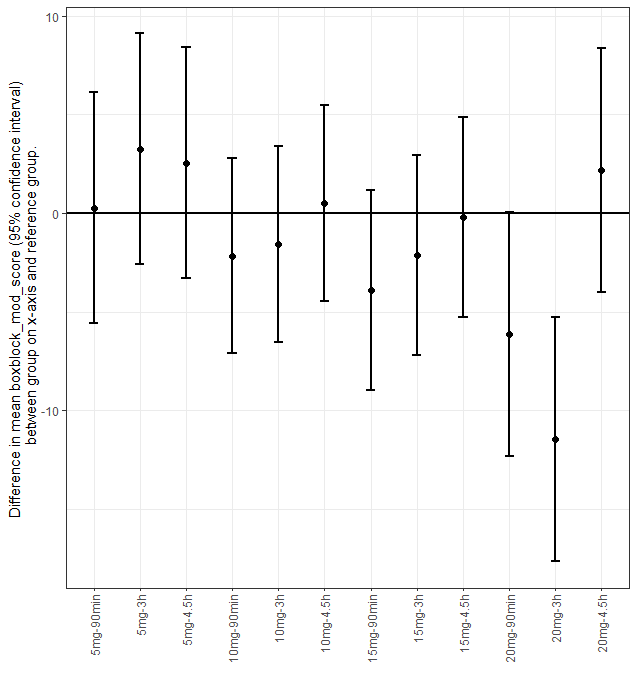

**Figure S5.4:** Caterpillar plot for boxblock_mod_score.

#### Digit Symbol Substitution Test

“dsst_score”

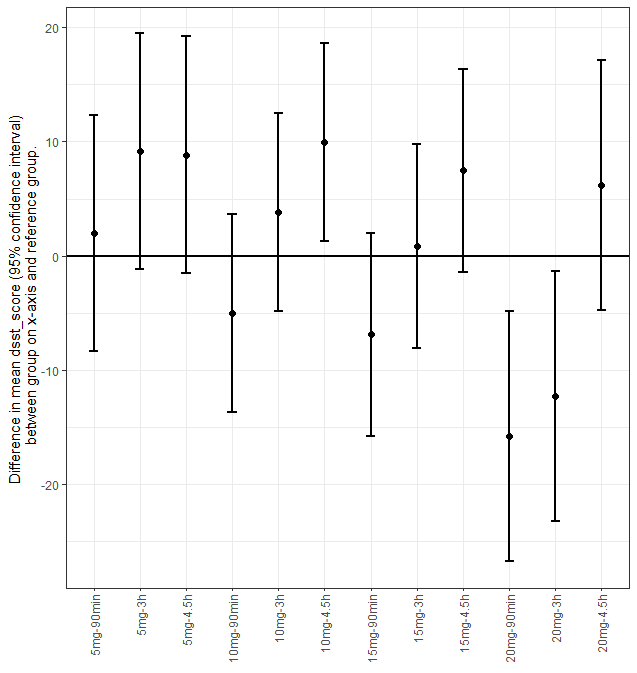

**Figure S5.5:** Caterpillar plot for dsst_score.

#### Reaction Time Ruler Drop Test (Mean)

“rt_ruler_mean”

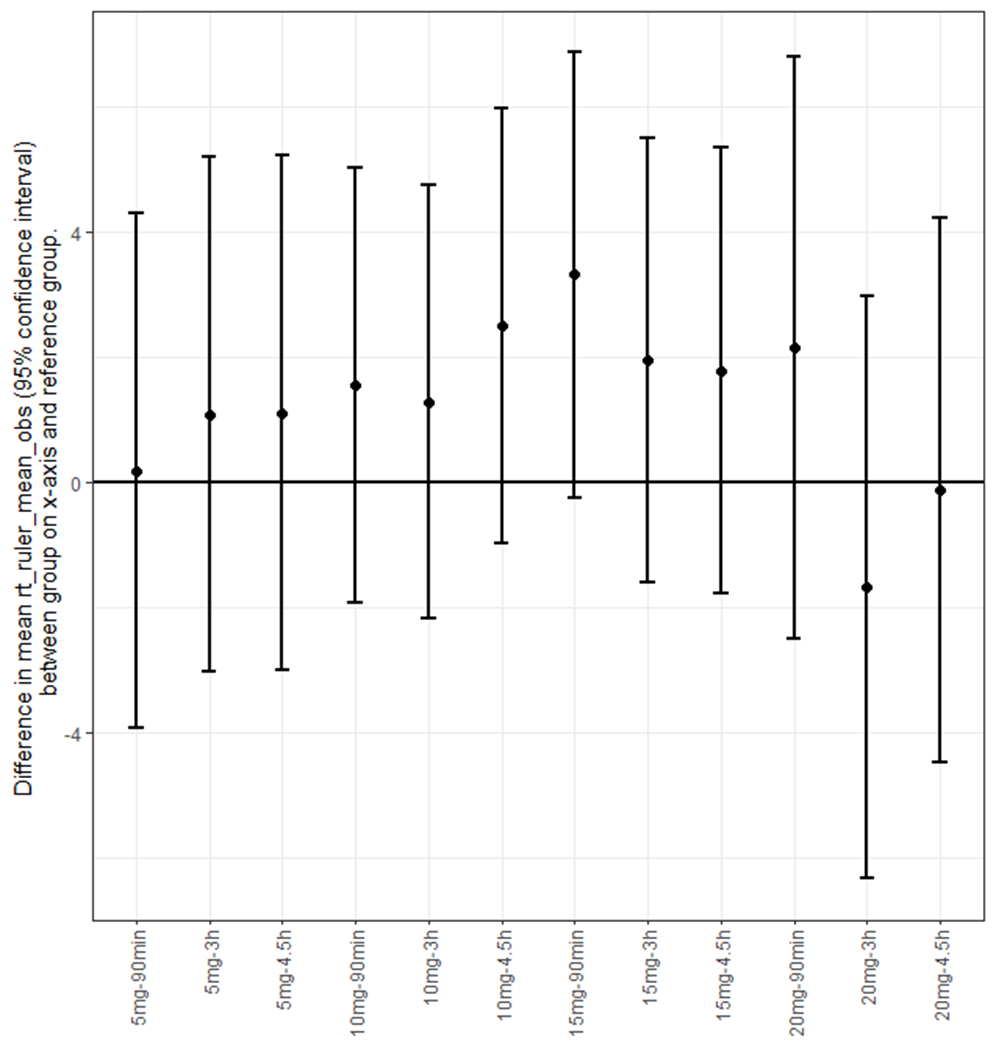

**Figure S5.6:** Caterpillar plot for rt_ruler_mean.

### Ego-Dissolution Inventory

#### Ego-Dissolution Inventory (Mean)

“edi_mean”

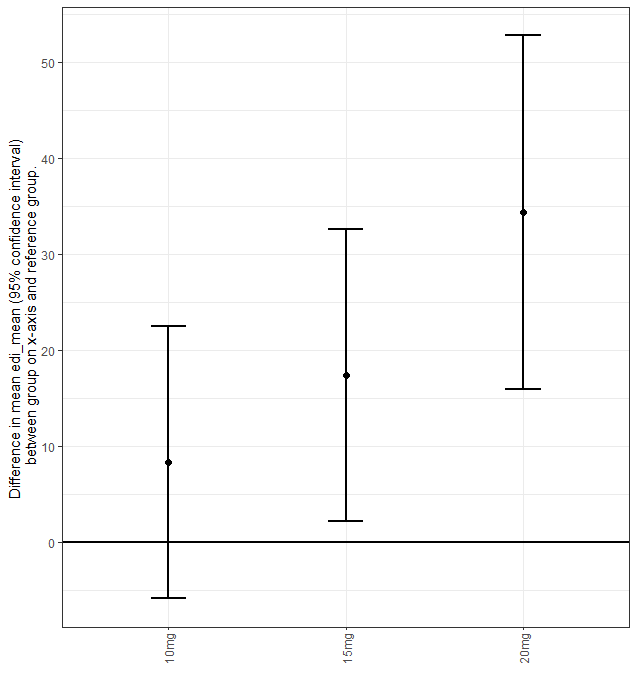

**Figure S5.7:** Caterpillar plot for edi_mean.

#### Q1: Dissolution of self

“edi_1”

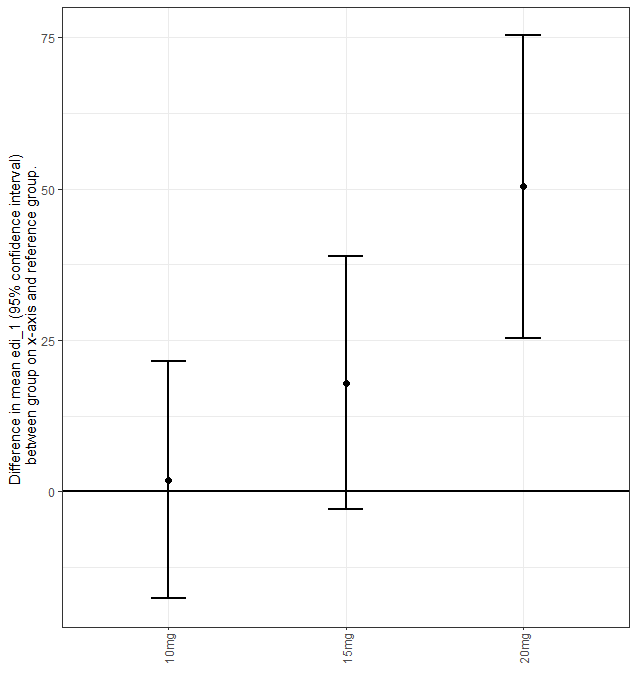

**Figure S5.8:** Caterpillar plot for edi_1.

#### Q2: Feeling one with the universe

“edi_2”

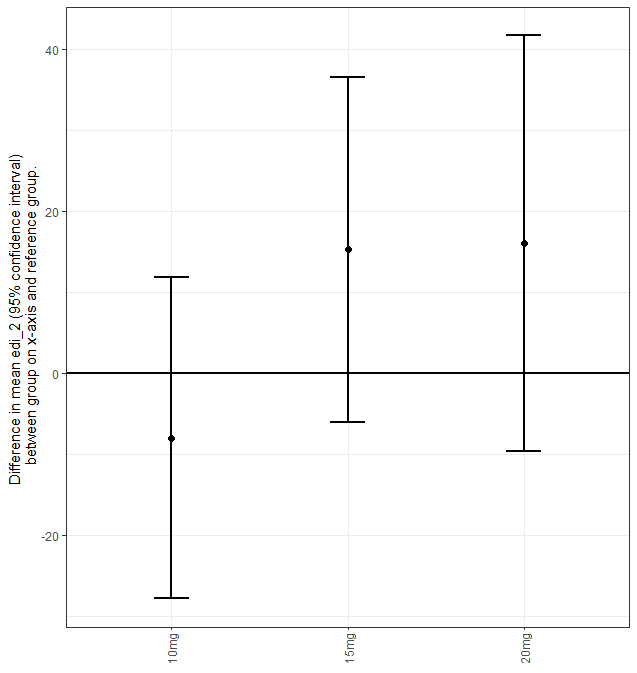

**Figure** **S5.9:** Caterpillar plot for edi_2.

#### Q3: Sense of union with others

“edi_3”

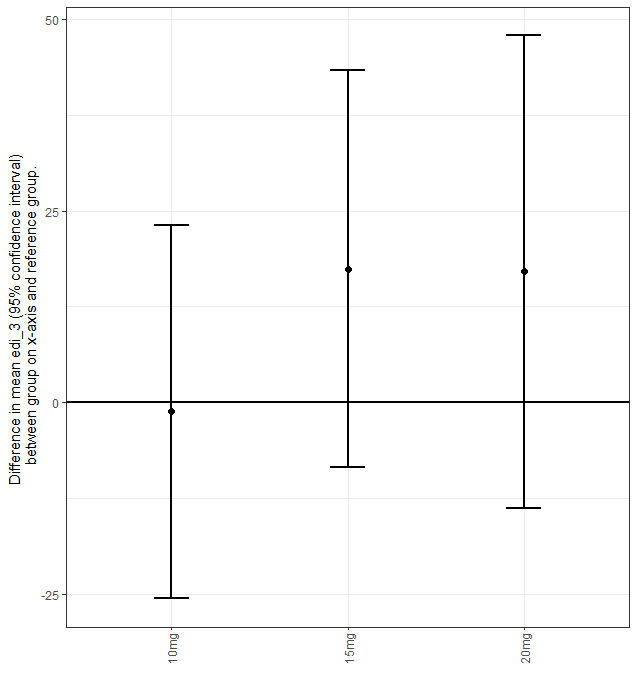

**Figure S5.10:** Caterpillar plot for edi_3.

#### Q4: Decrease in sense of self-importance

“edi_4”

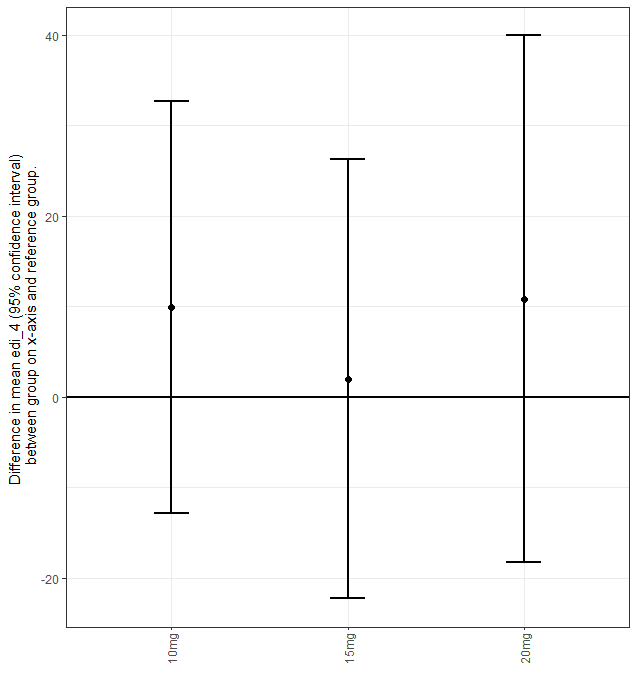

**Figure S5.11:** Caterpillar plot for edi_4.

#### Q5: Disintegration of self or ego

“edi_5”

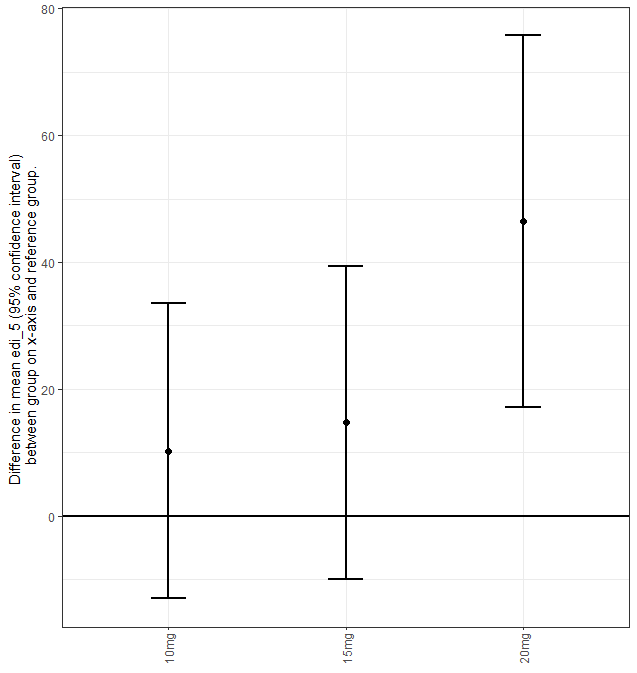

**Figure S5.12:** Caterpillar plot for edi_5.

#### Q6: Less absorbed with own issues/concerns

“edi_6”

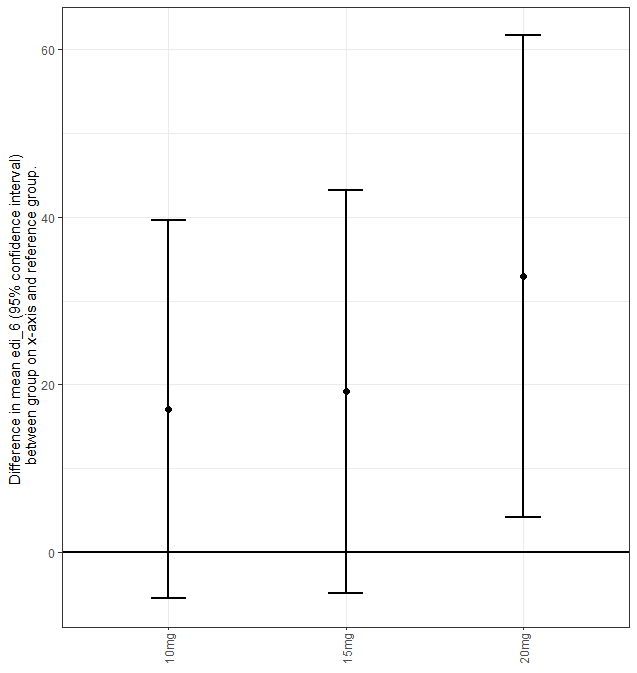

**Figure S5.13:** Caterpillar plot for edi_6.

#### Q7: Loss of sense of ego

“edi_7”

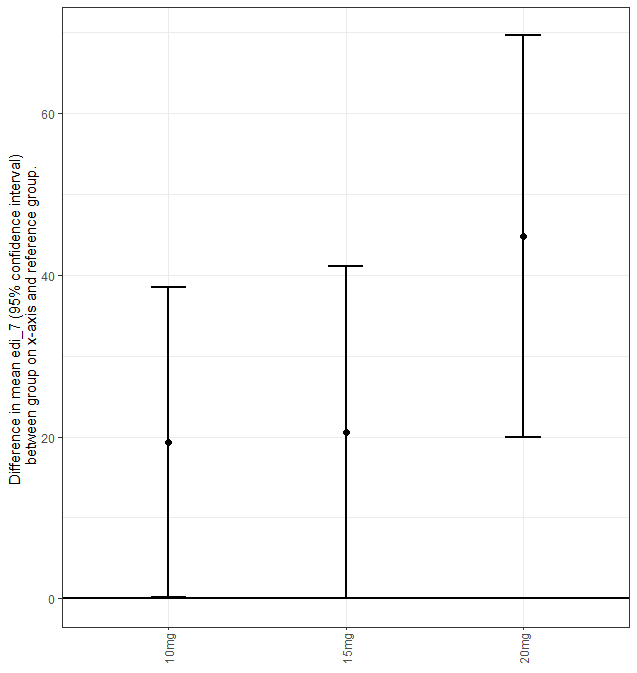

**Figure S5.14:** Caterpillar plot for edi_7.

#### Q8: All notion of self and identity dissolved away

“edi_8”

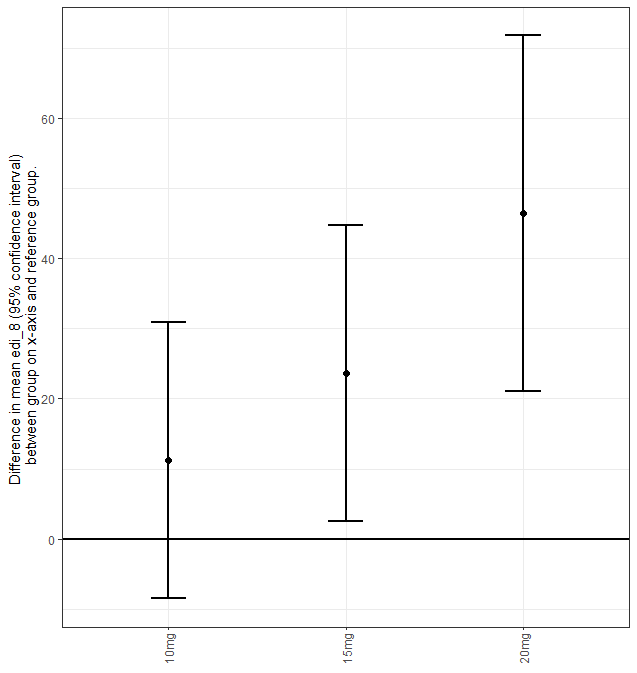

**Figure S5.15:** Caterpillar plot for edi_8.

### 5-Dimensional Altered States of Consciousness

#### 5-Dimensional Altered States of Consciousness (Mean)

“d5_asc_mean”

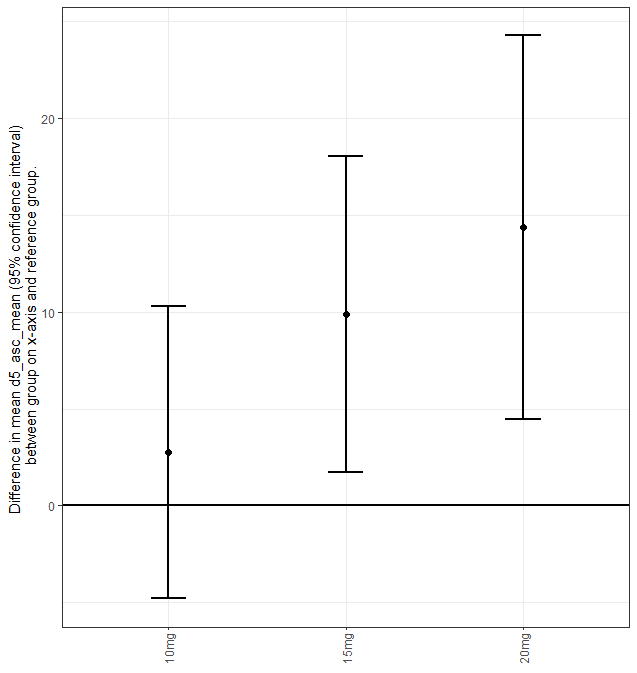

**Figure S5.16:** Caterpillar plot for d5_asc_mean.

#### Experience of Unity

“Experience_of_Unity”

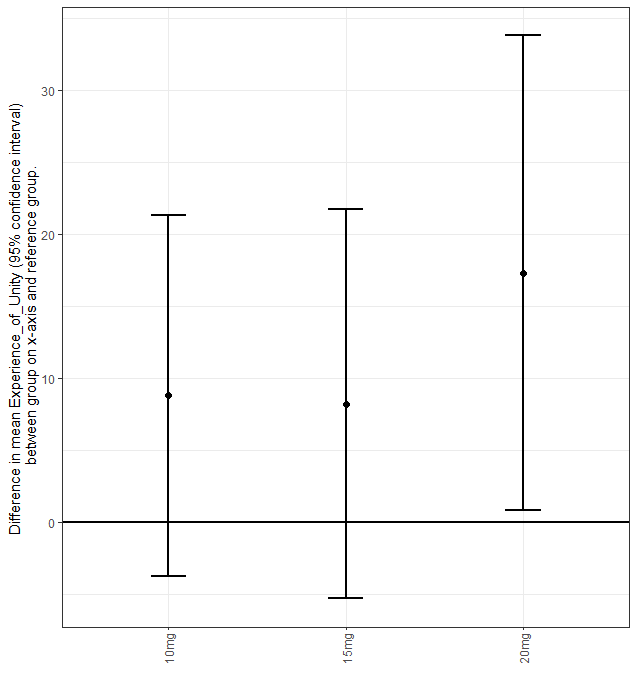

**Figure S5.17:** Caterpillar plot for Experience_of_Unity.

#### Spiritual Experience

“Spiritual_Experience”

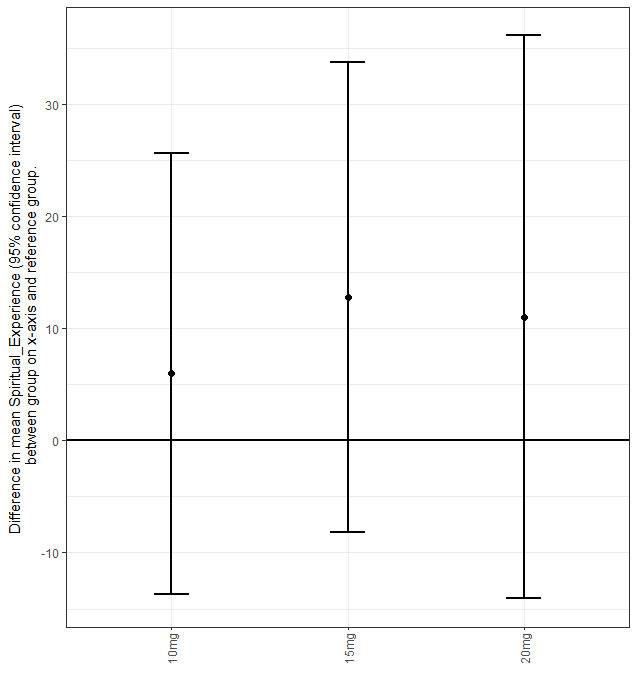

**Figure S5.18:** Caterpillar plot for Spiritual_Experience.

#### Blissful State

“Blissful_State”

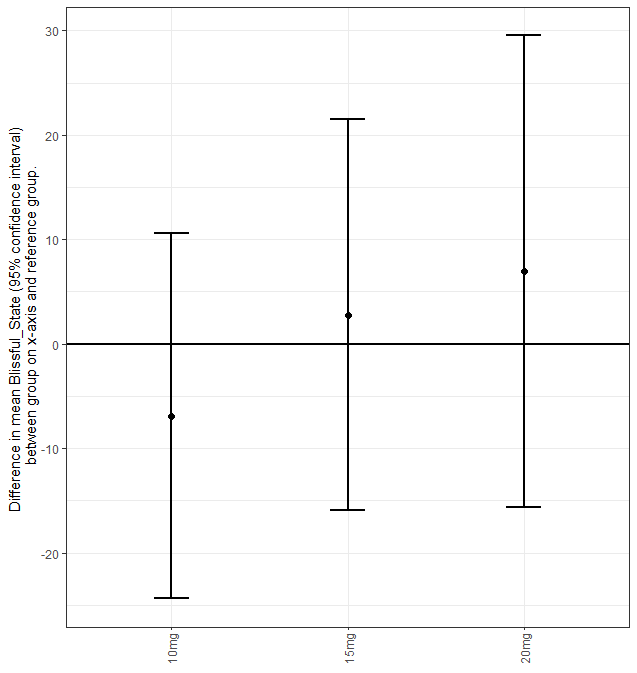

**Figure S5.19:** Caterpillar plot for Blissful_State.

#### Insightfulness

“Insightfulness”

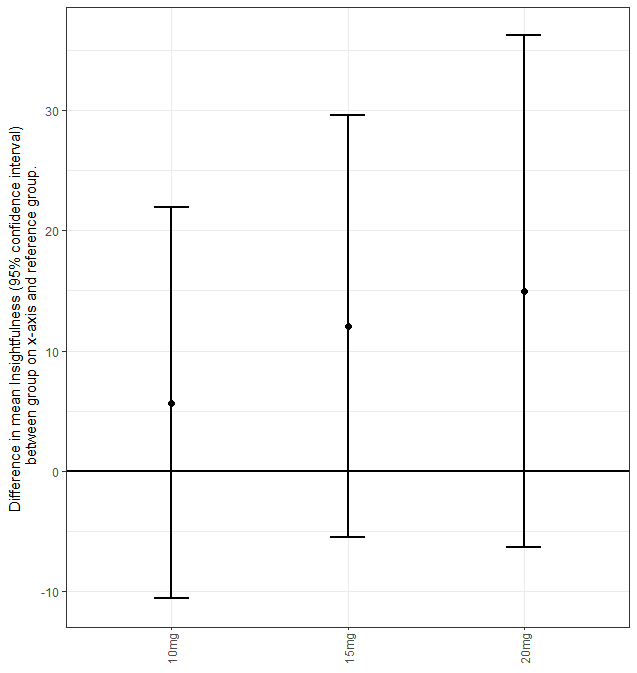

**Figure S5.20:** Caterpillar plot for Insightfulness.

#### Disembodiment

“Disembodiment”

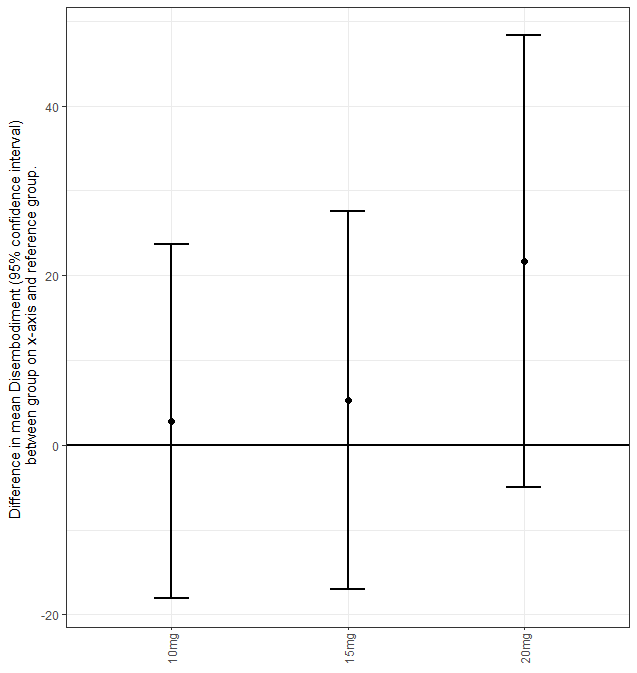

**Figure S5.21:** Caterpillar plot for Disembodiment.

#### Impaired Control and Cognition

“Impaired_Control_and_Cognition”

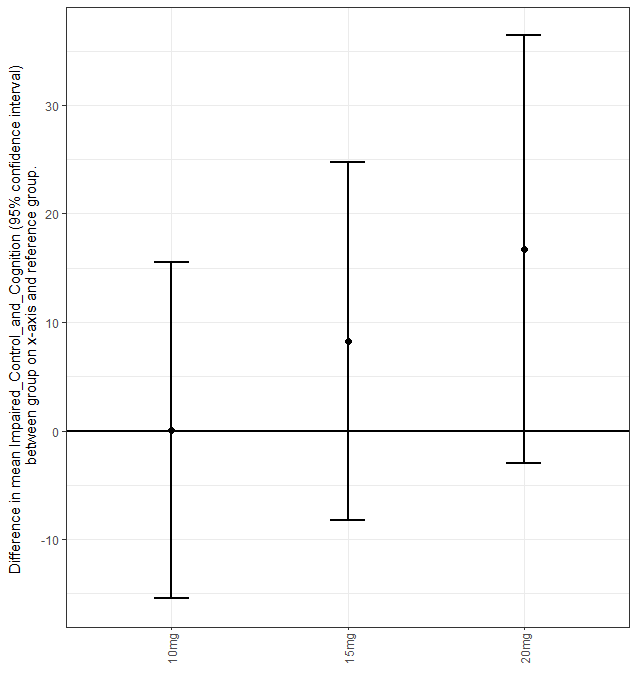

**Figure S5.22:** Caterpillar plot for Impaired_Control_and_Cognition.

#### Anxiety

“Anxiety”

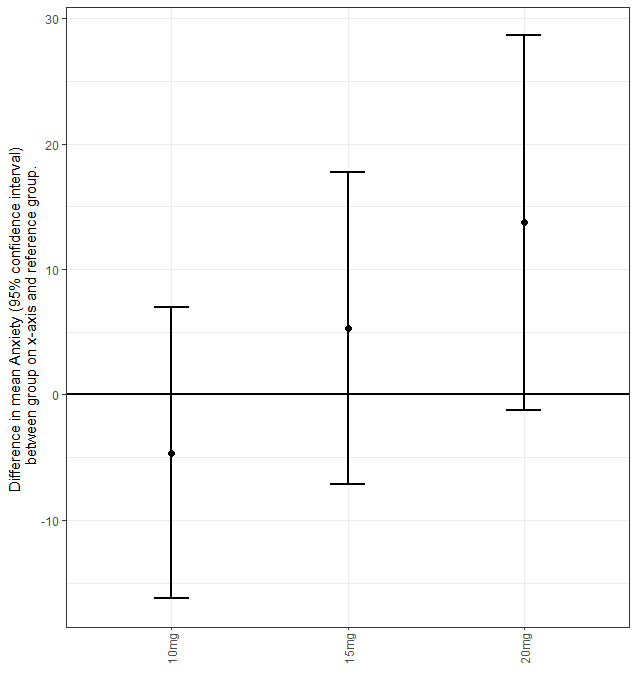

**Figure S5.23:** Caterpillar plot for Anxiety.

#### Complex Imagery

“Complex_Imagery”

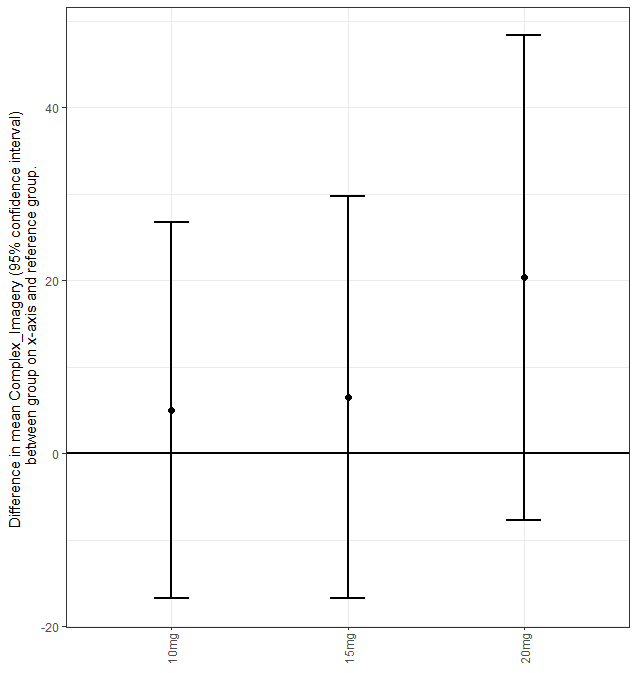

**Figure S5.24:** Caterpillar plot for Complex_Imagery.

#### Elementary Imagery

“Elementary_Imagery”

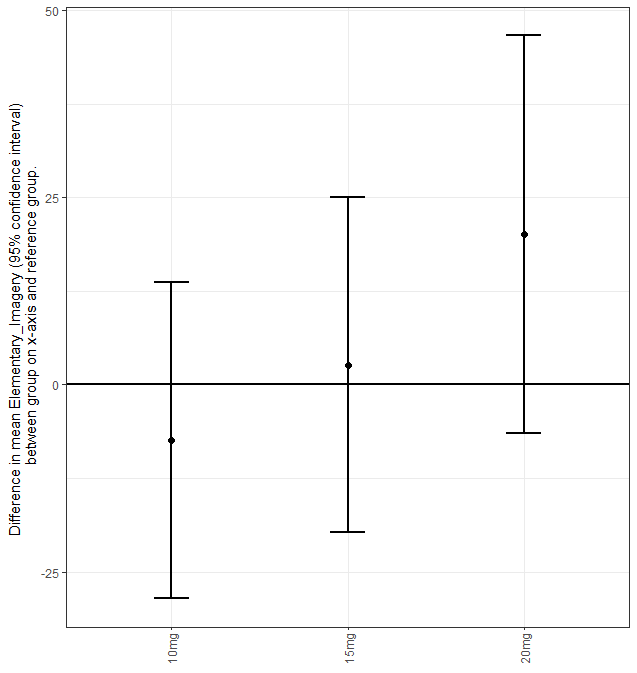

**Figure S5.25:** Caterpillar plot for Elementary_Imagery.

#### Audio Visual Synesthesiae

“Audio_Visual_Synesthesiae”

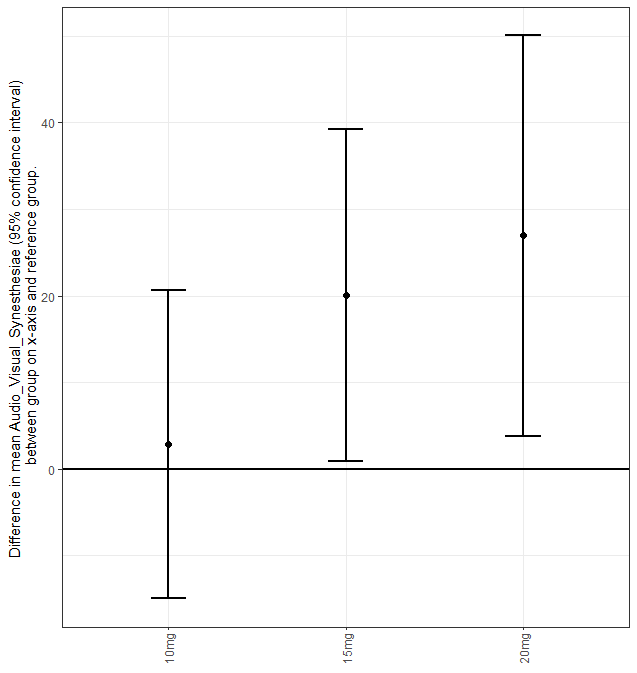

**Figure S5.26:** Caterpillar plot for Audio_Visual_Synesthesiae.

#### Changed Meaning of Percepts

“Changed_Meaning_of_Percepts”

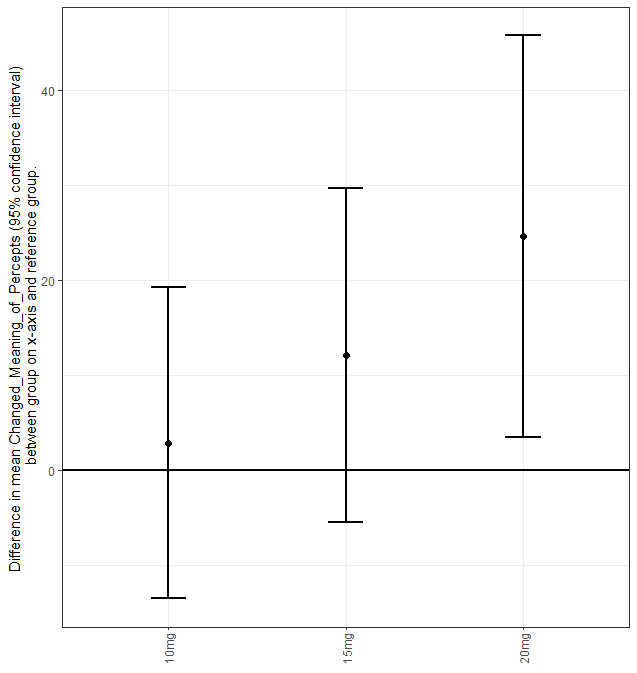

**Figure S5.27:** Caterpillar plot for Changed_Meaning_of_Percepts.

### Administrative Information

**Author Names:** Dr Chiranth Bhagavan, Professor Olivia Carter, Dr Glenn Nielsen, Professor David Berlowitz, Ms Sara Issak, Associate Professor Sabine Braat, Dr Sophie Zaloumis, Mr Zachary Attard, Ms Gina Oliver, Ms Deanne Mayne, Dr James Rucker, Dr Matthew Butler, Dr Orwa Dandash, Dr Alexander Bryson, and Professor Richard A. Kanaan.

**Corresponding Author:** Dr Chiranth Bhagavan, Department of Psychiatry, University of Melbourne, Austin Health, 145 Studley Rd, Heidelberg VIC 3084, Australia;; phone: +61 493 766 597.
