## Supplementary Information 5 for "A Randomised, Triple-Blind, Dose-Finding Study of the Impact of Psilocybin on Motor Function in Healthy Participants"

### Supplementary Information 1

### Statistical Analysis Plan

| **Authors:** | A/Prof Sabine Braat, University of Melbourne  (statistician) |
| --- | --- |
|  | Dr Sophie Zaloumis, University of Melbourne  (statistician) |
|  | Dr Chiranth Bhagavan, University of Melbourne  (clinical lead) |
| **SAP Version/Date:** | Version 1 (before unblinding)  (date: August 18, 2024)  Version 2 (after unblinding)  (date: September 1, 2025) |
| **Protocol version:** | Version 5.1  (date: May 16, 2024) |

1 Introduction

Healthy participants will be recruited on a volunteer basis for the dose finding study of psilocybin-assisted therapy. Each healthy participant will receive three doses of psilocybin, according to a Williams study design. Recruited participants will either take 5mg, 10mg and 15mg or 10mg, 15mg and 20mg. Each dose will be separated by at least 1 week and the order of treatment sequence will be randomised and blinded for each participant.

A planned sample size of 12 participants has been chosen for the following reasons:

- This is a realistic number of healthy participants that can be recruited over a 3-month period.
- This number of healthy participants is likely to be sufficient to determine if they are capable of completing physiotherapy after each of the three psilocybin dosing sessions (see Figure 1).
- This allows for an equal number of participants for each possible treatment dose sequence of the Williams design. While carryover effects between each dose are not anticipated, a Williams design allows for balance of first order carryover effects.

More details are described in the study protocol and published protocol paper [1].

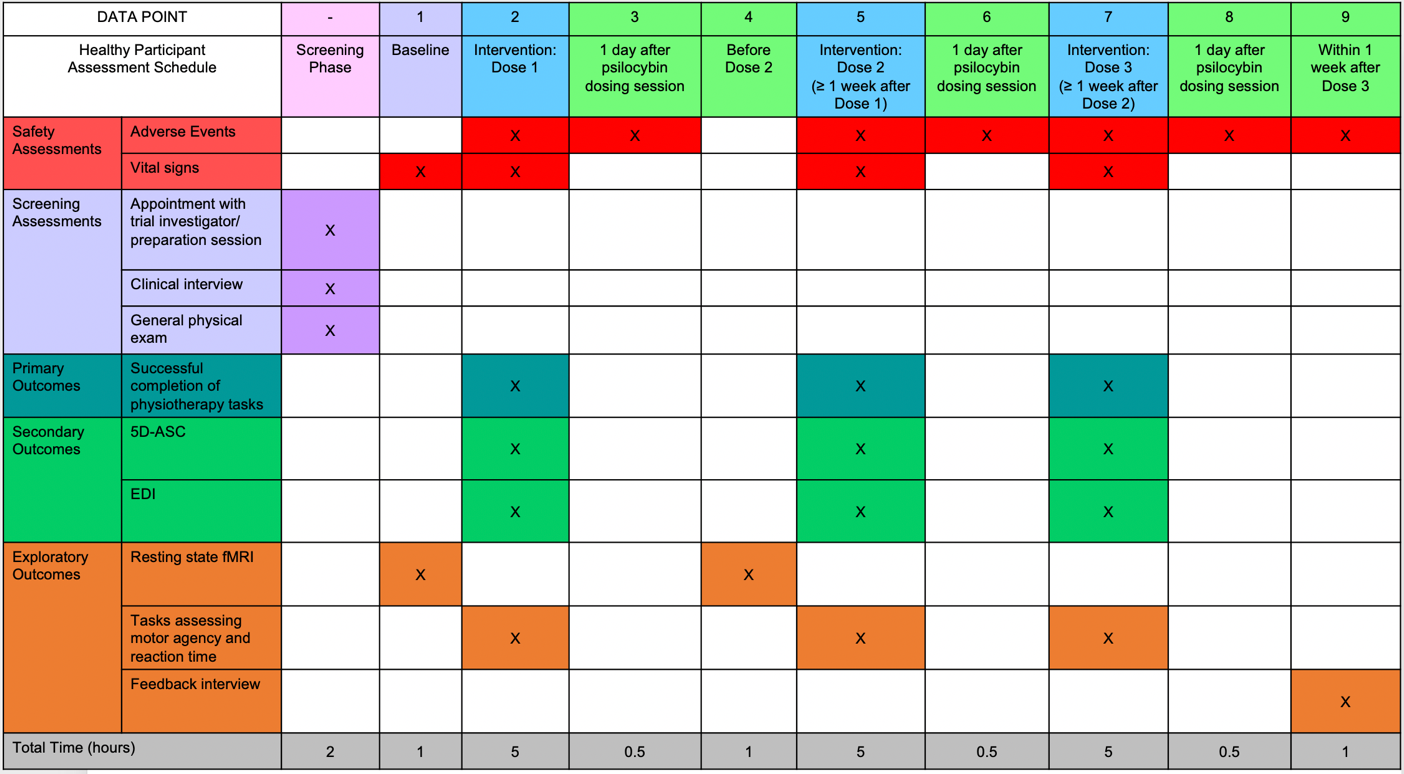

**Figure 1	Healthy Participant Assessment Schedule**

2 Analysis Objectives

**Primary aims**

1. To assess the feasibility of performing movement tasks during the acute effects of low-to-moderate doses of psilocybin.
2. To inform the maximum dose of psilocybin, up to 20 mg (inclusive), at which healthy participants can successfully complete a series of movement tasks during the acute drug effects.
3. To evaluate the safety of performing movement tasks during the acute effects of low-to-moderate doses of psilocybin.

**Exploratory aims**

1. To explore the effects of low-to-moderate doses of psilocybin on additional domains of motor function.
2. To explore the effects of low-to-moderate doses of psilocybin on verbal fluency.
3. To explore the subjective intensity of low-to-moderate doses of psilocybin.
4. To explore the effects of low-to-moderate doses of psilocybin on resting-state-derived measures of brain activity.
5. To explore the qualitative effects of low-to-moderate doses of psilocybin and performing movement and cognitive tasks during the acute drug effects.

3 Analysis sets/Populations

All available data of all healthy participants who meet the below eligibility criteria will be included in the analysis set, irrespective of study completion. No data after withdrawal of consent will be included.

**Inclusion criteria**

Eligible healthy participants for the dose-finding study will be:

- Adults aged 18 to 65 years
- No history of FND
- Who have volunteered for the study
- Capacity to provide informed consent

**Exclusion criteria**

All participants will be screened on all exclusion criteria.

*Medical Exclusion Criteria:*

- Cardiovascular conditions: poorly-controlled hypertension, angina, ischemic heart disease, a clinically significant ECG abnormality (e.g. atrial fibrillation), transient ischemic attack (TIA), stroke, peripheral or pulmonary vascular disease (no active claudication)
- A diagnosis of epilepsy or previous seizures
- A diagnosis of dementia
- A history of Chronic Kidney Disease or Chronic Liver Disease
- Known conditions putting the participant at risk for hypercalcaemia, Cushing's syndrome, hypoglycaemia, syndrome of inappropriate antidiuretic hormone secretion, or carcinoid syndrome
- Insulin-dependent diabetes; if taking oral hypoglycaemic agents, the participant is only excluded if they also have a history of hypoglycaemia
- Females who are pregnant, nursing or trying to conceive
- Use of medications contraindicated with psilocybin, that are inappropriate to cease for the necessary time period before/after the dosing session
- Patients enrolled in another clinical trial involving an investigational product

*Psychological Exclusion Criteria:*

- Current or previous diagnosis of any psychotic disorder, including Schizophrenia, Schizoaffective Disorder, Schizophreniform Disorder, Brief Psychotic Disorder, Delusional Disorder, Schizotypal Personality Disorder, Substance/Medication Induced Psychotic Disorder or Psychotic Disorder due to another medical condition
- Current or previous diagnosis of Bipolar I or II disorder
- First degree relative with diagnosed Schizophrenia, Psychotic Disorder, or Bipolar I or II Disorder
- A history of attempted suicide or mania
- Current or previous diagnosis of substance use disorder (excluding caffeine and nicotine)
- Previous regular use, or current use of psychedelic agents
- Current diagnosis of other psychiatric conditions judged to be incompatible with safe exposure to psilocybin (as determined by research staff)

4 Endpoints

**Participant characteristics and disposition**

- Includes age, gender, country of birth, height and weight
- Study completion
- Protocol deviations

**Primary outcomes**

- Combined total (range 0-26) of the de Morton Mobility Index (DEMMI) raw total score (range 0-19) and Functional Neurological disorder (FND) extension module (a.k.a. Functional Movement Exploration (FME)) total score (range 0-7). Higher scores are indicative of better performance
- Adverse and serious adverse events
- Vital signs

**Exploratory outcomes**

- Additional recordings related to DEMMI and FND extension module:
  - DEMMI duration
  - DEMMI total score (range 0-100), defined as the sum of its individual items. Higher scores are indicative of better performance
  - FND extension module total score (a.k.a. FME) (range 0-7), defined as the sum of its individual items. Higher scores are indicative of better performance
  - Time participant could hold tandem stand with eyes closed for (>10 < 30 seconds)
  - Time participant could stand on one leg (up to 30 seconds)
  - Number of errors in the verbal task (months in year)
  - Number of repetitions in 15 seconds (each arm) in the finger to nose test
- ~~Verbal fluency (prior to dosing and at the peak of the psilocybin effect [~ 2 hours after dosing]):~~
  - ~~Phonemic verbal fluency (generation of words from initial letters within a pre-set time of 60 seconds). Higher scores are indicative of better performance~~
  - ~~Semantic verbal fluency (generation of words from a given category within a pre-set time of 60 seconds). Higher scores are indicative of better performance~~
- Action Research Arm Test (ARAT):
  - ARAT total score (range 0-57), defined as the sum of all 19 items each scored using a 4-point ordinal scale (range 0-3). Higher scores are indicative of better performance
  - ARAT sub-scores for 4 subscales, defined as grasp (sum of 6 items, range 0-18), grip (sum of 4 items, range 0-12), pinch (sum of 6 items, range 0-18), and gross movement (sum of 3 items, range 0-9). Higher scores are indicative of better performance
- Box & Block Test:
  - Original version: Participants are scored based on the number of blocks transferred from one compartment to the other compartment in 60 seconds with the dominant hand. Higher scores are indicative of better manual dexterity
  - Modified version: The standard version is repeated with a shield between the participant’s vision and their hands and a mirror to view their movements via reflection. Higher scores are indicative of better manual dexterity
- Ruler drop:
  - Reaction time (seconds) for dominant hand. Lower scores are indicative of better performance
- Digit Symbol Substitution Test (DSST):
  - Number of symbols correct in 90 seconds using dominant hand. Higher scores are indicative of better performance
- Blinding efficacy:
  - Participant dose guess
  - Trial physiotherapist dose guess
- Ego-dissolution inventory (EDI):
  - EDI total score (range 0-100), defined as the mean score of all 8 items each scored on a visual analogue scale with the following statements at the lower and upper end, respectively: “No, not more than usually” (0=minimum) and “Yes, I experienced this completely/entirely.” (100=maximum). Higher scores are indicative of greater ego dissolution
- 5 Dimensional Altered States of Consciousness (5D-ASC):
  - Consists of 94 questions scored on a visual analogue scale (VAS) from 0 to 100 with the following statements at the lower and upper end, respectively: “No, not more than usually” and “Yes, much more than usually.”.
  - 11 dimensions (see Appendix 1 for details) (0 = minimum to 100 = maximum):
    - Experience of Unity
    - Spiritual Experience
    - Blissful State
    - Insightfulness
    - Disembodiment
    - Impaired Control and Cognition
    - Anxiety
    - Complex Imagery
    - Elementary Imagery
    - Audio-Visual Synesthesia
    - Changed Meanings of Percepts

Higher scores are indicative of greater altered states of consciousness

Data collected using fMRI, video footage (to assess movement quality), and qualitative interviews is outside of the scope of this SAP.

5 Handling of Missing Values and Other Data Conventions

Summary statistics will be based on available data, no missing data handling techniques will be applied. If an item or response is missing on a scale, then the total/ summary scale will be missing.

6 Statistical Methodology

Endpoints described in section 4 Endpoints will be summarised using descriptive statistics overall and by dose-level (baseline, 5 mg, 10 mg, 15 mg, and 20 mg) and time-point (if applicable). Change from baseline may also be derived for endpoints and summarised using descriptive statistics overall and by dose-level (5 mg, 10 mg, 15 mg, and 20 mg) and timepoint (if applicable). For some endpoints, summary statistics may be presented by block (low and high dose). Continuous variables will be summarized using n (non-missing sample size), mean, standard deviation, median, minimum, maximum, or range and categorical variables using counts and percentages (based on the non-missing sample size) of observed category levels.

**Participants characteristics and disposition**

Descriptive statistics for demographic and baseline variables will be presented overall and by dose-level.

The following will be reported to summarise participant disposition:

- Number of participants assessed for eligibility
- Number of participants not meeting the inclusion criteria and declined to participate
- Number of participants allocated to a dosing sequence
- Number of participants completing the study
- Number of participants not completed and reason
- Protocol deviations (number and type of protocol deviation)

**Primary outcomes**

If the available data permits, the primary analysis will be performed separately for each block (low and high dose). This will consist of a linear mixed effects model to compare the mean primary outcome (combined total of DEMMI and FND module scores) between dose-levels. The linear mixed effects model [2] will include the following terms:

- Dose-level (5 mg, 10 mg and 15 mg for low dose block and 10 mg, 15 mg, and 20 mg for high dose block)
- The primary outcome measured at the preparation session/baseline will be included as a covariate
- Dose session order (1^st^, 2^nd^ or 3^rd^) to capture secular time trends
- Random effect for participant. Captures the dependency between observations measured on the same participant
- Random effect for assessment time. Captures the dependency between observations measured in the same dosing session within a participant
- Residual error term capturing the difference between the observed primary outcome values and the predicted/fitted values from the model

The effect of the highest doses (Low dose block: 10 mg and 15 mg; High dose block: 15 mg and 20 mg) compared to the lowest dose (Low dose block: 5 mg; High dose block: 10 mg) will be presented as differences in means, 95% confidence intervals, and p-values.

If there’s little variation in the primary outcome, linear mixed effects modelling cannot be performed and only descriptive statistics for the primary outcome will be reported overall and by dose-level (baseline, 5 mg, 10 mg, 15 mg, and 20 mg) and time-point.

Participants experiencing adverse events and serious adverse events will be listed and the number and percentage of participants experiencing at least one adverse event and serious adverse event will may be summarised (if numbers allows) overall and by the dose group, severity, and relationship to investigational medicinal product. Descriptive statistics for vital signs will be presented overall and by dose-level and time-point.

**Exploratory outcomes**

Descriptive statistics will be presented overall, and by dose-level and time-point (if applicable). Results may be presented graphically (e.g. bar plots, line plots, spider plots [3]).

To assess blinding efficacy, cross tabulations of the following for each dosing session will be presented:

- Participant dose guess and actual dose received
- Trial physiotherapist dose guess and actual dose received

**Sensitivity analyses**

One participant with deafness was unable to conduct the verbal task (months in year) of the FND extension module in its original format and as a result the participant scored 0 (i.e., unable to complete) on this task. A sensitivity analysis of the primary outcome (combined total of DEMMI and FND module scores) and FND extension module score will be conducted whereby these total scores will be transformed into a percentage of the maximum total score (range 0-100) in addition to the total (sum) score. Descriptive statistics will be reported overall and by dose-level (baseline, 5 mg, 10 mg, 15 mg, and 20 mg) and time-point.

Some participants did not grasp the ruler in the ruler drop test and as a result the participant scored a missing value for that attempt. There is no standard convention regarding the scoring of such a result and the final score is averaged over the non-missing scores. A sensitivity analysis of the ruler drop will be conducted whereby the missing score will be set to the length of the ruler (30cm) plus 1cm before averaging across the three attempts. Descriptive statistics will be reported overall and by dose-level (baseline, 5 mg, 10 mg, 15 mg, and 20 mg) and time-point.

**Subgroups**

Output will be generated by gender (male, female).

7 Post-hoc changes performed after unblinding

Changes to the Sections 1-6 in Version 1 of the SAP before unblinding:

- Section 4 Endpoints: Analysis of verbal fluency data will be reported separately and is therefore out of scope of this SAP.

In addition to the plan prior to unblinding as outlines in Section 6 Statistical Methodology, the following post-hoc analyses were conducted.

*Motor function*

- Motor outcomes were analysed using a linear mixed effects model including the following terms:
  - Because dose-level and time after dose share a common baseline can only include a combined variable with the following categories: baseline, 5mg-90min, 5mg-3h, 5mg-4.5h, 10mg-90min, 10mg-3h, 10mg-4.5h, 15mg-90min, 15mg-3h, 15mg-4.5h, 20mg-90min, 20mg-3h and 20mg-4.5h
    - Categorical covariate
    - Fixed effect
    - Baseline is the reference group
    - Interpretation of parameter estimates: Difference in mean outcome at each dose-time after dose combination compared to baseline
  - Random effect for participant. Captures the dependency between observations measured on the same participant (i.e., each participant has multiple outcome values measured at each dose).
    - Makes sure standard errors and 95% confidence intervals for fixed effects account for the dependency between observations from the same participant
  - Residual error term capturing the difference between the observed primary outcome values and the individual predicted/fitted values from the model
- Note: rt_ruler_mean – currently treating any dropped or did not catch ruler results as missing

*EDI and 5D-ASC*

- EDI and the 11 dimensions of the 5D-ASC were analysed using a linear mixed effects model including the following terms:
  - Dose-level: 5 mg, 10 mg, 15 mg, and 20 mg
    - Categorical covariate
    - Fixed effect
    - 5 mg the reference group
    - Interpretation of parameter estimates: Difference in mean outcome for 10mg, 15mg or 20 mg compared to 5mg
  - Random effect for participant. Captures the dependency between observations measured on the same participant (i.e., each participant has an outcome value measured at some point after each dose).
    - Makes sure standard errors and 95% confidence intervals for fixed effects account for the dependency between observations from the same participant.
  - Residual error term capturing the difference between the observed primary outcome values and the individual predicted/fitted values from the model.

*Results/Outputs*

- For all post-hoc analysis the following results/outputs were presented:
  - Plot of observed individual profiles
  - Caterpillar plot showing the estimated mean difference and 95% confidence interval
  - Table of parameters estimates from linear mixed effects model
    - These are the values plotted in the caterpillar plot
  - Graphical assessment of model assumptions: linearity, constant (homogeneity) of variance, influential observations, normality of residuals, and normality of random effects
  - Plots comparing fitted (model derived) individual profiles and the estimated population average profile to the observed individual profiles

### Appendix 1 Scoring for the 5D-ASC

**11 dimensions**

1) Experience of Unity (average of 5 items)

18 - Everything seemed to unify into an oneness

34 - Felt one with my surroundings

41 - Experienced a touch of eternity

42 - Conflicts and contradictions seemed to dissolve

52 - Experienced past, present, and future as a oneness

2) Spiritual Experience (average of 3 items)

9 - Felt connected to a higher power

81 - Experienced a kind of awe

94 - My experience had religious aspects to it

3) Blissful State (average of 3 items)

12 - Experienced boundless pleasure

86 - Experienced profound inner peace

91 - Experienced an all-embracing love

4) Insightfulness (average of 3 items)

50 - Felt very profound

69 - Had insights into connections that had previously puzzled me

77 - Had very original thoughts

5) Disembodiment (average of 3 items)

26 - Felt as if I no longer had a body

62 - Had the impression I was out of my body

63 - Felt as if I was floating

6) Impaired Control and Cognition (average of 7 items)

8 - Felt like a puppet or marionette

27 - Felt incapable of making even the smallest decision

38 - Had difficulties in distinguishing important from unimportant

47 - Felt as if I were paralyzed

64 - Felt isolated from everything and everyone

67 - Was not able to complete a thought; my thoughts repeatedly became disconnected

78 - Had the feeling that I no longer had my own will

7) Anxiety (average of 6 items)

32 - Was afraid that the state I was in would last forever

43 - Was scared without knowing exactly why

44 - Experienced everything as frighteningly distorted

46 - Experienced my surroundings as strange and weird

56 - Felt threatened

89 - Had the feeling that something terrible was going to happen

8) Complex Imagery (average of 3 items)

39 - Saw whole scenes roll by with closed eyes or in complete darkness

72 - Could see images from my memory or imagination with extreme clarity

82 - My imagination was extremely vivid

9) Elementary Imagery (average of 3 items)

14 - Saw regular patterns with closed eyes or in complete darkness

22 - Saw colors with closed eyes or in complete darkness

33 - Saw brightness or flashes of light with closed eyes or in complete darkness

10) Audio-Visual Synesthesiae (average of 3 items)

20 - Sounds seemed to influence what I saw

23 - Shapes seemed to be changed by sounds or noises

75 - The colors of things seemed to be altered by sounds or noises

11) Changed Meaning of Percepts (average of 3 items)

28 - Some everyday things acquired special meaning

31 - Things in my environment had a new strange meaning

54 - Objects in my surroundings engaged me emotionally much more than usual

### Administrative Information

**Author Names:** Dr Chiranth Bhagavan, Professor Olivia Carter, Dr Glenn Nielsen, Professor David Berlowitz, Ms Sara Issak, Associate Professor Sabine Braat, Dr Sophie Zaloumis, Mr Zachary Attard, Ms Gina Oliver, Ms Deanne Mayne, Dr James Rucker, Dr Matthew Butler, Dr Orwa Dandash, Dr Alexander Bryson, and Professor Richard A. Kanaan.

**Corresponding Author:** Dr Chiranth Bhagavan, Department of Psychiatry, University of Melbourne, Austin Health, 145 Studley Rd, Heidelberg VIC 3084, Australia;; phone: +61 493 766 597.
